# The macroeconomic and epidemiological impacts of Covid-19 in Pakistan

**DOI:** 10.1101/2023.10.06.23296657

**Authors:** Henning Tarp Jensen, Marcus R. Keogh-Brown, Rosalind M. Eggo, Carl A. B. Pearson, Sergio Torres-Rueda, Maryam Huda, Muhammad Khalid, Wahaj Zulfiqar, CMMID COVID-19 Working Group, Richard D. Smith, Mark Jit, Anna Vassall

## Abstract

“Coronavirus Disease 2019” (C19) is a respiratory illness caused by “new Coronavirus” SARS-CoV-2. The C19 pandemic, which engulfed the world in 2021, also caused a national C19 epidemic in Pakistan, who responded with initial forced lockdowns (15-30 March 2020) and a subsequent switch to a smart lockdown strategy, and, by 31 December 2020, Pakistan had managed to limit confirmed cases and case fatalities to 482,506 (456 per 100,000) and 10,176 (4.8 per 100,000). The early switch to a smart lockdown strategy, and successful follow-up move to central coordination and effective communication and enforcement of Standard Operating Procedures, was motivated by a concern over how broad-based forced lockdowns would affect poor households and day-labour. The current study aims to investigate how the national Pakistan C19 epidemic would have unfolded under an uncontrolled baseline scenario and an alternative set of controlled non-pharmaceutical intervention (NPI) policy lockdown scenarios, including health and macroeconomic outcomes. We employ a dynamically-recursive version of the IFPRI Standard Computable General Equilibrium model framework (Lofgren, Lee Harris and Robinson 2002), and a, by now, well-established epidemiological transmission-dynamic model framework (Davies, Klepac et al 2020) using Pakistan-specific 5-year age-group contact matrices on four types of contact rates, including at home, at work, at school, and at other locations (Prem, Cook & Jit 2017), to characterize an uncontrolled spread of disease. Our simulation results indicate that an uncontrolled C19 epidemic, by itself, would have led to a 0.12% reduction in Pakistani GDP (−721mn USD), and a total of 0.65mn critically ill and 1.52mn severely ill C19 patients during 2020-21, while 405,000 Pakistani citizens would have lost their lives. Since the majority of case fatalities and symptomatic cases, respectively 345,000 and 35.9mn, would have occurred in 2020, the case fatality and confirmed case numbers, observed by 31. December 2020 represents an outcome which is far better than the alternative. Case fatalities by 31. December 2020 could possibly have been somewhat improved either via a more prolonged one-off 10 week forced lockdown (66% reduction) or a 1-month forced lockdown/2-months opening intermittent lockdown strategy (33% reduction), but both sets of strategies would have carried significant GDP costs in the order of 2.2%-6.2% of real GDP.

## Introduction

The world is currently undergoing the worst health crisis of the past century. While other communicable and non-communicable diseases may, in the past, have incurred larger disease burdens, the emergence and rapid spread of the novel Corona virus, SARS-CoV-2, and its fellow Corona Virus Disease 2019 or Covid-19 (C19), has put the entire world on high alert. The risk of overwhelmed health systems (at society-level), and lack of effective treatments or vaccines, led governments, around the world, to recognize the critical threat to public health, and the need to resort to sometimes draconian non-pharmaceutical interventions (NPIs). While many C19 vaccines were developed at break-neck speed, and the recent emergence of effective treatments, in the form of antiretroviral pills, have recently given hope to many around the world, the combination of delays in production and global roll-out of vaccination programmes, and the continued emergence of new and more infectious SARS-CoV-2 strains in multiple locations, means that complementary NPI restrictions are likely to continue to be needed, for quite some time, in countries around the world, and especially in large and potentially vulnerable low- and middle-income countries (LMICs) such as Pakistan.

During the first 1½ years, the C19 epicentre has shifted from East Asia to Europe, to North and South America, South Asia, and, more recently, back to Europe. Outside American and European epicentres, most Asian nations fared better, until exponential growth took hold in early 2021, following a range of super spreader events including religious festivals and political rallies (Reuters 2021o), and the emergence of the new B1617 delta variant of concern (WHO 2021c, WHO 2021e), However, in spite of the severity of India’s massive delta variant corona-wave this year, including 451,435 confirmed case fatalities by 14. October 2021 (Ritchie 2021) and with reports of excess mortality exceeding official numbers by as much as an order of magnitude (Anand, Sandefur & Subramanian 2021), other populous Asian nations, including Indonesia and, in particular, Pakistan, have, in spite of rising clinical outcomes, been more effective in containing their national epidemics, and, in the case of Pakistan, have, so far, avoided exponential growth and kept trend growth under relative control (Ritchie 2021).

As of 14 October 2021, Pakistan had recorded 1,261,685 cases and 28,201 case fatalities related to C19 (Ritchie 2021). In spite of the promise of vaccination programs, which started to be rolled-out globally in December 2020 and early 2021, access to vaccines at population scale has proven to be an issue for a populous lower-middle-income country like Pakistan. While Pakistan has been assisted by China and, more recently the US, in their clinical C19-response, and signed-up for UN’s COVAX facility (VOA 2021, WHO 2021b), and, in spite of being involved in Phase 3-trials of the Chinese CanSinoBIO Ad5-nCoV vaccine (DW 2020, WHO 2020b) which subsequently has led to a (limited) local vaccine packaging production of Ad5-nCoV vaccines locally referred to as PakVac (Reuters 2021q), the Pakistani government only made their first official announcement, on 31. December 2020, of plans to purchase 1.2mn doses of the Chinese Sinopharm BBIBP-CorV vaccine (Reuters 2020a) and followed by slow public roll-out due to worries, among prioritized healthcare workers (HCWs), about the efficacy and safety of BBIBP-CorV (HPW 2021); And, while the first COVAX-vaccine delivery of 1.2mn was received on 8 May 2021 (Reuters 2021l). A Gallup Pakistan poll from November 2020 further suggested vaccine scepticism is high, and that as many as 37% of Pakistanis may not want to get a vaccine once one becomes available (Reuters 2020b). In spite of more recent success in accessing vaccine deliveries and handling vaccine roll-out, with 50mn vaccine doses delivered on 28 August (DAWN 2021e), and 100mn doses delivered on 24 October 2021 (DAWN 2021f), Pakistani authorities will therefore, most likely, need to make controlled achievement of herd immunity a medium-to-long-term policy goal, also because many Pakistanis do not come in for their second booster jab (DAWN 2021g) and because signs were starting to emerge, in November 2021, of problems in reaching remote rural citizens without formal forms of identification (DAWN 2021h). In the meantime, NPIs will need to remain a corner-stone of Pakistani C19 epidemic control due to the threat of new variants, underscored by the experience of the March-May SL lockdown period, following B117 alpha variant spreading and becoming dominant throughout Pakistan in May 2021 (Reuters 2021m)^1^and covering the Eid ul Fitr holidays in May (DAWN 2021d), and the July-September SL lockdown, during the Eid ul Azha and Pakistan independence Day holidays and following B1617 delta variant simultaneously being reported dominant in Karachi/Sindh on 15 July (DAWN 2021i) and in Lahore/Punjab on 17 July (DAWN 2021j).

Lockdowns have been instituted four times, in Pakistan, over the past year, including March-May and November-December 2020, and March-May and July-September 2021. The spring 2020 lockdown included: (1) Phase 1: Forced lockdown (FL) period (15-30. March 2020), (2) Phase 2: Partial Lockdown (PL) period (31. March – 5. April 2020), and (3) Phase 3: Smart Lockdown (SL) period (6-15. April 2020) (Rukh, Nafees & Khan 2020). The Pakistani authorities quickly decided against FL-style NPIs, in favour of SL-style NPIs, striking a balance between “Standard Operating Procedure” (SOP) information campaigns, and alternative SL-style measures such as tele-schooling and one-day-a-week/two-day-a-week business lockdowns, limited to virus hotspots, and only under exceptional circumstances, general bans on public rallies, gatherings, and indoor weddings, and closure of entertainment venues such as cinemas and theatres (Statesman 2020). Nationwide lockdown has been vehemently resisted by federal authorities as evidence by federal resistance to a 10-day general lockdown of Sindh province including the Karachi – referred to Pakistani politicians as the “country’s economic lifeline” (DAWN 2021k).

The aim of the Pakistani SL-style NPI measures has been to reduce human activity and contact numbers, and more recently to provide incentives for getting immunized, on the one hand, and to minimize income and poverty impacts on the other, but no assessment has so far been made of the relative economic costs and health benefits of implementing alternative FL-style NPI strategies in Pakistan. This study aims to fill this gap by constructing and applying a Pakistan Computable General Equilibrium (CGE) model, based on the “Standard model” framework (Löfgren, Lee Harris & Robinson 2002), with province-level households, to assess the relative economic costs and health benefits of pursuing alternative one-off and intermittent FL-style NPI strategies, in Pakistan, and thereby provide a backdrop, for Pakistani policymakers, to assess the relative merits and relative success of their preferred SL-style strategy, and to provide an upper bound of the potential value of vaccine roll-out to Pakistani society as measured by the GDP cost of alternative FL-style NPI control strategies. Epidemiological scenarios are simulated using a state-of-the-art C19 epidemiological model for Pakistan (Pearson et al. 2020; Davies, Klepac et al. 2020b).

The rest of the paper is organized as follows: The Background section will discuss Pakistan’s C19 epidemic(s), both at the national and provincial levels, and put Pakistan’s experiences with C19 into regional and global context; The Methods section will present our combined methodological framework for epidemiological and macroeconomic modelling of C19, and also present our uncontrolled C19 baseline scenario and our three NPI-focussed policy scenarios; The following Results section will present and interpret our results and put them into a Pakistan context; And the Conclusion and Discussion section will highlight our main conclusions, and discuss caveats to our results and avenues for future work.

## Background

Pakistan is a federal country, which, since the adoption of the 25^th^ constitutional amendment, passed in 2018, has consisted of seven administrative units, including four provinces: Balochistan, Khyber Pakhtunkhwa (KPK), Punjab, and Sindh; and three territories including: Azad Jammu and Kashmir (AJK), Gilgit-Baltistan (GB), and Islamabad Capital Territory (ICT).

### Disease background

C19 is a respiratory illness which is caused by SARS-CoV-2, and which spreads via facial droplets or via contact between hands/objects and the face. The basic reproduction number (R0) has generally been considered to be high (≈2.5-3.5). Whilst Pakistan-specific estimates initially indicated that it may ‘only’ have been around 1.6-1.9 (Uddin et al 2020; Mustafa et al 2020; Ali, Imran & Khan 2020; Ullah & Khan 2020), this is still twice as infectious as seasonal flu (R0 around 1.4), implying that vaccine-induced herd immunity against the original strain would have required coverage levels of 50-60% with a highly efficacious vaccine (or >70% with efficacy levels around 60%).^2^ However, the repeated outbreaks of variants of concern with elevated transmissibility, this year, implies that required coverage levels could well approach 80%^3^ (or higher with less efficacious vaccines), something which could be a matter of serious concern especially in light of reported Pakistani vaccine-scepticism (Reuters 2020b).

The morbidity of C19 cases varies from asymptomatic and mild “common cold”-type symptoms, to severe pneumonia, acute respiratory failure, and, for groups-at-risk, high rates of mortality (Zhou et al 2020). Case fatality rates among unvaccinated population groups range from 1:50,000 among children at age 10, and 1:10,000 among young adults at age 25, to 1:70 among retirees around age 65, and 1:7 among elderly around age 85 (Levin et al 2020), and, for Pakistani C19 cases requiring hospitalization, illness progression seems to be characterized by an initial asymptomatic stage of approx. 5 days followed, for severe cases, by approx. 8 days of hospitalization in general ward, and, for critical cases, by approx. 10 days of hospitalization, including around 1/3 of the time in general ward and 2/3 of the time in intensive care, until death or discharge (Torres-Rueda et al 2020).

Evidence has also emerged on long-term health sequelae among recovered C19 cases, referred to as “Long-COVID”. Among recovered C19 inpatients, critical cases may have significantly increased risks of long-term health sequelae at six months after discharge, including diffusion impairment, anxiety/depression, and fatigue/muscle weakness (Huang et al 2021), and SARS and MERS data indicate that the reduced diffusion capacity may affect 11–45% of hospitalized C19 patients at 12 months after discharge (Ahmed, Patel et al 2020) and may persist for as long as 7 to 15 years (Ahmed, Patel et al 2020; Zhang et al 2020). A small-scale Pakistani study of recovered C19 in- and outpatients confirmed that the majority of cases experienced fatigue, poor sleep quality, anxiety, and dyspnea, almost half experienced a loss of taste and smell regardless of severity of illness, three-quarters of severe cases experienced severe problems in daily activities, and they also found evidence of pulmonary disease even among mild cases (Iqbal et al 2021). Since these indications about Long-Covid manifestations, and negative health system capacity impacts during peak epidemic periods, are still being studied and mapped, they will not be included in our simulation analyses.

### National Pakistan disease background^4^

Since gaining its independence in 1947, Pakistan has experienced rapid population growth, but, with declining fertility rates over the recent decade, the end of the demographic transition is finally in sight. Nonetheless, Pakistan still has a fairly young population, with 81% of the population aged 0-39 and 3.4% aged 65+ (2019) (GBD 2020). A recent study found the size of the 65+ age group to be a significant predictor of relative C19 caseloads in a cross-section of 182 countries (Nguiemkeu & Tadedjeu 2021), something which may help explain Pakistan’s relatively low C19 counts, which on 14 October 2021, included 595 confirmed cases and 13.3 case fatalities per 100,000 population, as compared with approx. 10,000-13,500 confirmed cases and approx. 180-220 case fatalities per 100,000 population in other populous countries such as US, Brazil, Mexico^5^, UK, Italy, France, and Spain (Ritchie 2021).

Other factors identified by Nguiemkeu & Tadedjeu as significantly affecting relative C19 caseloads, include demographic and geographic variables such as population density (+), urban population share (+), and temperature (−). In terms of population density, Pakistan’s rapid post-independence population growth has increased the population density from 59.8 people/km2 in 1961 to 275.3 people/km2 in 2018, a change from being the 84^th^ to 47^th^ most densely populated country in the world (WB 2020). At the same time, disease burdens of major infectious diseases such as TB, HIV/AIDS, and malaria have been trending downwards over recent years (GBD 2020), indicating that improvements in sanitation and other preventive public health measures have, so far, allowed Pakistan to maintain control over the spread of communicable disease.

In terms of climatic conditions, the climatic variations between Pakistani provinces and territories do not seem to have created systematic variations in C19-epidemics between the regions of Pakistan. After quickly gaining control over their epidemics during June-July 2020, the four provinces experienced accelerating transmission in late-October 2020, suggesting that seasonality, rather than climate, has been the main driver of transmission during 2020 (Figure 1). Variants of concern, perhaps together with holiday gatherings, have driven the Corona waves during 2021, but, in spite of upward-sloping regional transmission curves, all provinces and territories have so far, by early-November 2021, managed to keep their C19-epidemics under relative control, perhaps with the exception of the urbanized and densely populated ICT, which is showing the highest regional confirmed case count and case fatality rates amounting to respectively 3,092 confirmed cases and 28.3 deaths per 100,000 population per 14 October 2021 (Table 1), but which was also the first major urban center to report partial resident vaccine coverage of 70 % on 6 September 2021 (DAWN 2021l) and 84% on 18 October 2021 (DAWN 2021m).

**Figure 1.**
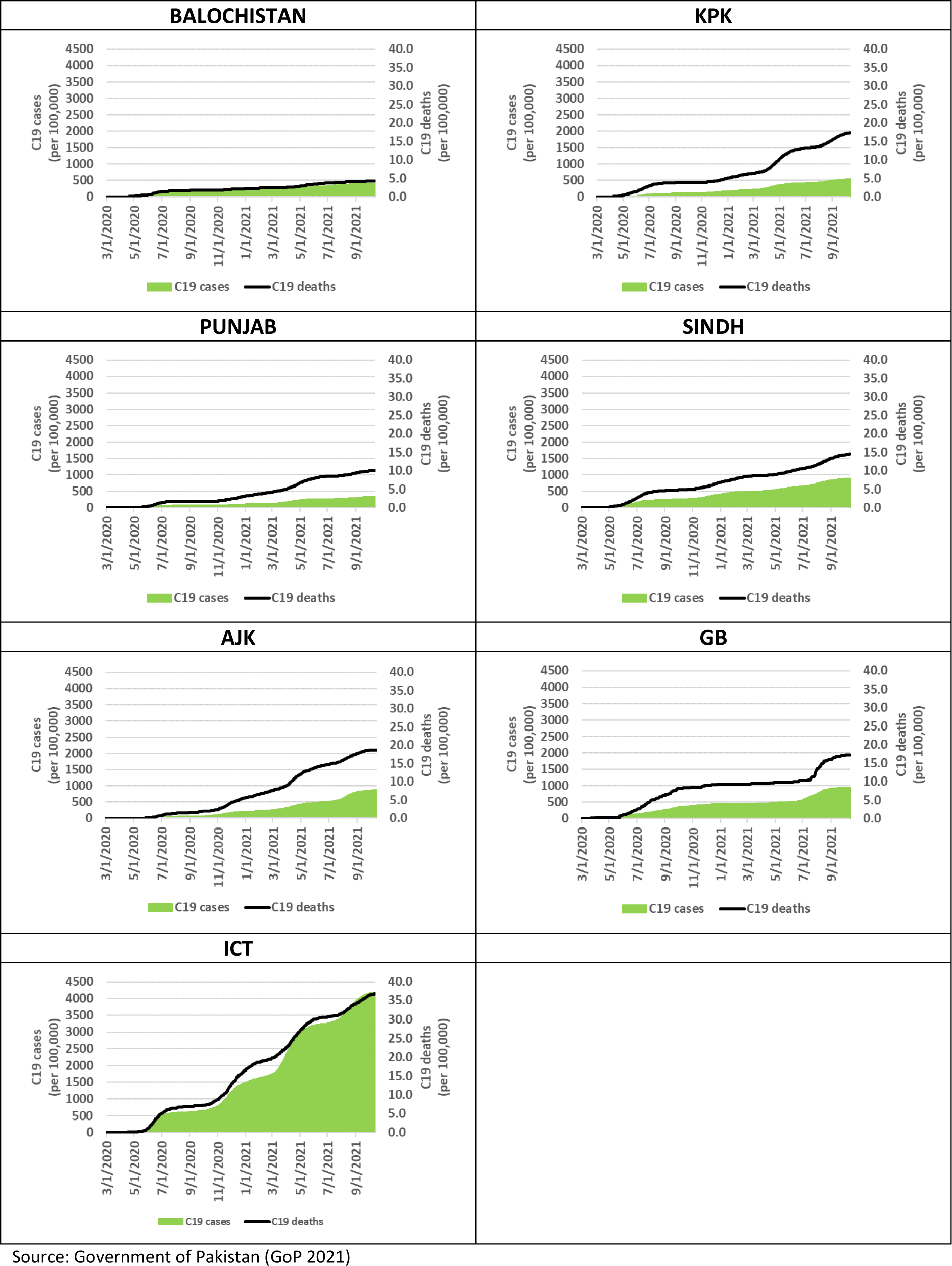
Pakistan C19 clinical outcomes by Province & Territory 1. March 2020 – 14. October 2021 (per 100,000 population)

**Table 1.** Confirmed C19 cases, case fatalities and CFRs (by 14 October 2021)

|  | Confirmed case fatalities<br>(per 100,000 population) | Confirmed cases<br>(per 100,000 population) | Case Fatality Rates<br>(per confirmed case) |
| --- | --- | --- | --- |
| Balochistan | 3.08 | 289.4 | 1.06% |
| KPK | 11.22 | 383.7 | 2.93% |
| Punjab | 7.23 | 253.3 | 2.85% |
| Sindh | 9.20 | 568.2 | 1.62% |
| AJK | 12.88 | 456.8 | 2.82% |
| GB | 9.88 | 499.2 | 1.98% |
| ICT | 28.26 | 3091.7 | 0.91% |

Source: own calculations based on Government of Pakistan (GoP 2021)

In the populous Punjab and Sindh provinces, while smart lockdown NPIs are likely to have played a key part in their epidemic containment, the climate may also have favoured out-door activities during at least the first part of the epidemic, until night-time temperatures started dropping below 20C towards end-of-October 2020. More recently, due to the highly infectious B117 having become dominant throughout Pakistan (Reuters 2021m), 7-day transmission growth rates (of confirmed cases) in KPK and Punjab provinces, and ICT and AJK territories, accelerated from 1.9-2.5% end-of-February to peak at 7.3%-8.5% during the week ending 10 April, and subsequently to decelerate to 3.4-3.8% during the week ending 11 May (GoP 2021). During the same period, the two remaining provinces, Balochistan and Sindh, and the remaining territory, GB, experienced gradually increasing transmission, implying that all regions experienced relatively constant 7-day transmission growth rates of 1.2-3.9% during the week ending 11 May (ibid.) While geographically focussed FL-style NPI measures have been employed in Punjab (DAWN 2021a, 2021b), the Pakistan government continued to oppose national lockdowns by end-of-March (Reuters 2021i), but national guidelines on lockdowns has subsequently started to emerge, most recently in relation to the Eid holidays (DAWN 2021d). The reversal of Pakistani policy has come during a period of exponential transmission in neighbouring India (Figure B.4 in Appendix B) – an experience which has made India the new global C19 hotspot, and which has brought the horrors of laissez-faire policy to the attention of the world (Reuters 2021o).

### Pakistan vaccination programme and roll-out

The Drug Regulatory Authority of Pakistan (DRAP) had, by 31. March, granted emergency use authorisation to at least five vaccines including Sinopharm’s BBIBP-CorV vaccine (Reuters 2021c), AstraZeneca’s ChAdOx1 vaccine (Reuters 2021d), Gamaleya’s Gam-COVID-Vac “Sputnik V” vaccine (Reuters 2021e), CansinoBio’ Ad5-nCoV vaccine (Reuters 2021f), and the Sinovac vaccine (ET 2021). Interim/final phase 3 trial-evidence has indicated efficacy rates of 92% for the Gam-COVID-Vac (Logunov 2021) and 62% for ChAdOx1 (Voysey et al 2021b). No peer-reviewed evidence exists for the remaining vaccines, but news reports suggest efficacy rates of 79% for BBIBP-CorV (BioWorld 2020)^6^ and 51-84% for Sinovac (Reuters 2021p). In terms of efficacy against variants of concern, while no peer-reviewed evidence exists on the efficacy of Gam-COVID-Vac or BBIBP-CorV against the B117, B1351, and P1 variants, the evidence-base for ChAdOx1 is more developed. Hence, non-peer-reviewed working paper evidence indicates that ChAdOx1 may be almost as efficacious against B117-induced C19 as against wildtype-induced C19 (Emary et al 2021), while peer-reviewed evidence, conversely, suggests that it is not efficacious against the B1351 variant (Madhi et al 2021).

Pakistan’s vaccine roll-out, which started immediately upon receipt of the first batch of BBIBP-CorV on 1 February 2021, was initially delayed by worries, among prioritized healthcare workers (HCWs), about the efficacy and safety of BBIBP-CorV (HPW 2021)^7^ and by delayed COVAX deliveries^8^. In the context of rising B117 transmission, the DRAP, on 4 March, reversed their previous decision and approved BBIBP-CorV and Gam-COVID-Vac for the over-60s (NewsInt 2021), paving the way for expanding their vaccination campaign to non-HCWs with people prioritized in reverse order by age (BS 2021) – and the first 4,219 persons, in the prioritized age groups, were inoculated on 10-11 March 2021 (SAMAA 2021).

The government has previously announced that they are not planning to buy vaccines, but instead “…aims to tackle the COVID-19 challenge through herd immunity and donated vaccines from friendly countries like China” (HT 2021). That being said, Pakistani authorities continue to be focussed on the planned COVAX-related procurement of ChAdOx1/Covishield from the Serum Institute of India, with initially planned delivery of the first batch by the middle of March (delivered on 8 May (Reuters 2021l)) and a second larger delivery in June (ibid.). Moreover, they have recently turned their attention to harnessing the power of private markets. On 17. March 2021, the first private imports of 50,000 doses of Gam-COVID-Vac vaccines arrived in Pakistan (Devex 2021), and additional large-scale bulk shipments of Cansino’s Ad5-nCoV vaccines were expected to arrive in mid-April (Reuters 2021f, DAWN 2021c) but had not yet arrived in early-May 2021. Instead, the Pakistani authorities now expect shipments of 13mn doses of BBIBP-CorV, Ad5-nCoV, and Sinovac vaccines to arrive before end-of-June (Reuters 2021n). Regardless of the Pakistani authorities’ mixed signals, and in spite of their efforts to harness the power of private markets, vaccination roll-out seems likely to be slowed by restricted access to vaccine supplies, and the Pakistani government will therefore, most likely, need to make controlled achievement of herd immunity a medium-to-long-term goal.

### Macroeconomic modelling background

The macroeconomic C19 literature generally focuses on C19 as a health shock with economic repercussions (1) directly via health-related impacts and (2) indirectly via private and public mitigating actions. Research into the economics of C19 has, to a large degree, focused more on high-income countries (HICs) than on LMICs. However, some comprehensive monographs have emerged covering both global, HIC and LMIC perspectives (Baldwin & di Mauro 2020), and economists from international institutions have attempted to summarize the limitations which LMICs are facing in terms of structural limitations and potential policy options going forward (Loaysa & Pennings 2020).

A number of macroeconomic studies have emerged which focus on combined epidemiological and macroeconomic modelling of C19 illness and C19 NPI policy shocks. The group of aggregate Dynamic Stochastic General Equilibrium (DSGE) model studies, with endogenous epidemiological models, is focussed on aggregate social planning problems and optimal control strategies. In contrast, single-country and global Computable General Equilibrium (CGE) studies, are focussed on more detailed economic burden and NPI policy assessments; and, for one of the CGE studies (Keogh-Brown et al 2020) and one non-DSGE social planning-type framework (Haw et al 2020a, 2020b), focus is also on detailed transmission-dynamic epidemiological modelling of clinical health outcomes.

The group of aggregate DSGE intertemporal model studies employ simple but fully integrated Susceptible-Infected-Recovered SIR-type epidemiological models (Atkeson 2020a, 2020b). They also specify explicit social planning problems and use these to measure optimal lockdown strategies for minimizing macroeconomic and social welfare costs (Alvarez, Argente & Lippi 2020; Eichenbaum, Trebelo & Trabandt 2020; Jones, Philippon & Venkateswaran 2020a, 2020b). Drawbacks include limited sector-level detail, simple epidemiological model specifications with a typical lack of age-specific heterogeneity of contact patterns and clinical health outcomes, a lack of nuance in specification of lockdown strategies (typically focusing on the business closure aspect of lockdown strategies), and application of widely varying Value of Statistical Life (VSL) measures of mortality-related welfare losses (Alvarez, Argente & Lippi 2020, Jones, Philippon & Venkateswaran 2020a, 2020b). A key strength of the social planning DSGE approach is that it has managed to explicitly model private prevention behaviour, and model two key externalities including (1) the infection externality (each individual only focussed on own rather than society protection leading to undersupply of transmission-reducing behavioural change), and (2) the health system congestion externality (each individual not taking risk of health system congestion into account, again leading to undersupply of transmission-reducing behavioural change), and drawing attention to the scope for (US) tele-working to mitigate against the need for costly business closure lockdowns (Jones, Philippon & Venkateswaran 2020). While opportunities for tele-working may be more limited for Pakistan and other LMICs, the above results are suggestive and, for example, tele-health and tele-schooling initiatives have also been rolled-out in Pakistan (Farooq et al 2020).

The group of C19-focussed CGE model studies, currently in the public domain, includes one application of a single-country CGE model framework similar to the one applied here (Keogh-Brown et al 2020), and applications of various global model frameworks, including the ImpactECON Supply-Chain Model, a standard CGE model which is a modified version of the Global Trade Analysis Project (GTAP) model (Walmsley, Rose & Wei 2020), and the G-Cubed model, a hybrid DSGE/CGE model with aggregate intertemporal DSGE features, but with an underlying multi-sector CGE model which solves for annual equilibria over a very long 2015-2100 time horizon (McKibbin & Fernando 2020). Among the global studies, Walmsley et al employs a fairly simplistic specification of shocks, including only mandatory business closures of one and four month durations, while McKibbin and Fernando, in spite of being published very early (29 February 2020) and using scant data including 2003 SARS data from China and rough indicators for inferring country-specific shocks, goes much further and designs global sets of sophisticated country-specific scenarios involving direct and indirect health-related shocks (morbidity and mortality-related labour force impacts), equity risk premia, sector-specific (transport) costs of production, and consumer preference impacts. Public health costs of clinical health outcomes are, however, not inferred or analysed, and lockdown measures are not analysed either, presumably due to the early publication date. Notably, the McKibbin-study finds that “Hong Kong Flu”- and “Spanish Flu”-type pandemics would lead to health-related costs equivalent to respectively 2% and 8% declines in global GDP ($2.3 & $9.2 trillion), while (2) the Walmsley-study finds that four months of non-essential business closures would lead to a 21.6% decline of U.S. GDP ($4.6 trillion) and a 23.0% drop in employment (36.1 million unemployed workers).

Using the same “Standard Model” single-country methodology as the current study (see Methods section below), a UK single-country study analysed C19 lockdown scenarios covering a range of business closure NPIs (BC-NPIs), school closure NPIs (SC-NPIs), and home quarantine NPIs (HQ-NPIs) (Keogh-Brown et al 2020), derived from the widely recognized Imperial College COVID-19 Response Team’s preferred intermittent 2-month lockdown/1-month opening-up scenario (Ferguson, Laydon et al. 2020). A key strength of the single country CGE framework, in a health context, is its ability to accommodate exogenous epidemiological model predictions, and thereby allow for model-consistent public health assessments of economic and clinical health burdens, for non-communicable diseases (NCDs) (Jensen et al 2019a) and communicable diseases (CDs)(Keogh-Brown et al 2020), but without making assumptions about government preferences. Hence, the focus on analysing policy scenarios, rather than specifying social planning problems, allow decisions on trade-offs to be left up to policy makers – and, by the same token, policymakers are free to apply their own preferences when facing trade-offs between relative economic and clinical health impacts from various competing NPI policy strategies.

Finally, it should be mentioned that Imperial College has established a social planning-type framework where an epidemiological model is linked with an economic module via a social planning objective which, in this case, is real GDP (Haw et al 2020a, 2020b). The macroeconomic module is, however, rudimentary, consisting purely of fixed sector-specific production vectors, which are scaled by sector-specific BC-NPI lockdown shares, but which does not involve economic behaviour.

## Methods

We utilize a twin set of C19 epidemiological and macroeconomic models to analyse the health and economic burdens associated with (1) an uncontrolled C19 baseline, and (2) three alternative NPI-controlled policy scenarios in Pakistan during 2020-21. We apply the epidemiological model as a satellite model, and use it to produce health-related scenario-specific shocks, which we impose on the macroeconomic model to measure overall epidemiological and macroeconomic outcomes of our unmitigated baseline, and three policy scenarios.

Our dynamically-recursive macroeconomic model is specified around a core static macroeconomic CGE model framework, also known as the ‘Standard Model’ (Löfgren, Lee Harris & Robinson 2002) – a so-called multi-sector model which allows for analysing a multitude of production activities and retail commodities. Producers aim to maximize profits and consumers aim to maximize welfare, the government collects taxes to fund their recurring spending (mainly on essential public services), savings are collected and channelled into productive investment projects, and domestic retailers engage with foreign traders, to trade in import and export goods. In the current context of modelling NPI policy scenarios, the multi-sectoral nature of our framework has the added advantage that we can specify our model with a level of sectoral detail which allows for sensible disaggregation of the economy’s production sectors into essential and non-essential sectors (see appendix A for sector classifications).

We calibrated our static CGE model from a 2014 Pakistan Social Accounting Matrix (SAM) which was derived from the Global Trade Analysis Project (GTAP) database version 10 (GTAP 2019). The original 65 production activities were aggregated to 51 (see appendix A) with the twin aims to facilitate model simulations, but also to maintain sufficient detail to allow for separation of essential and non-essential production sectors, which is critical for specification and modelling of BC-NPIs within the macroeconomic model. Furthermore, we disaggregated the single SAM household account into four provincial representative households, in order to capture province-specific C19 epidemics and NPI scenarios, and also to allow for analyses of economic impacts across the four Pakistani provinces.^9^ Provincial household income and expenditure patterns were derived from the 2015-16 Household Integrated Income and Consumption Survey (HIICS) (PBS 2017) and used to disaggregate the household account, and the resulting unbalanced SAM matrix was balanced using standard cross minimum entropy techniques (Golan, Judge, & Robinson 1994; Robinson, Cattaneo & El-Said 2001).

Household-specific factor ownership data and sector-specific employment data (including numbers of skilled and unskilled workers belonging to the four provincial households and employed in each of the 51 production sectors) were also extracted from the 2015-16 HIICS survey (PBS 2017). The modelling of province-level labour factor ownerships, within the macroeconomic model, was critical as it allowed us to model shocks to province-specific effective labour factor participation in response to health-related C19 labour absenteeism and labour supply losses due to employed case fatalities. Furthermore, in order to model epidemic progression over 2020-21, we turned the model into a recursively-dynamic model by adding labour and capital factor-updating equations.

The labour factor updating equations were calibrated by producing a set of province-level demographic projections, based on a standard demographic model specification (Jensen et al 2019b). The demographic model was calibrated to a set of UN population projections (UN 2020) and disaggregated based on WorldPop province-level demographic data (WorldPop 2020), and age- and gender-specific labour force participation rates (PBS 2018) were subsequently applied to produce a complete set of baseline province-level labour factor ownership growth path projections for 2014-2021. We also extracted time series of capital stocks and capital depreciation rates for Pakistan from the Penn World Tables data base version 9.1 (PWT 2020) and used these data to initialize and calibrate our capital updating equation. With the dynamically-recursive model completed, including fully specified factor-updating equations, we used historical Pakistan GDP growth rates (WB 2020) to establish 2020 as the base year for our 2020-21 policy simulations, and we, subsequently, calibrated a counterfactual 2020-21 ‘non-C19’ counterfactual growth path to historical real (2.8% p.a.) and nominal (5.2% p.a.) GDP growth rates from the period 2015-18 (ibid.) against which the C19 baseline and NPI policy scenario impacts could be assessed (For details on CGE model parametrization and model closure, see appendix C).

Given that C19 is a contact-transmissible disease, we utilize a well-established transmission-dynamic framework, initially fitted to the epidemic in Wuhan, and based on globally estimated age-varying susceptibility to SARS-CoV-2 virus infection and age-varying C19 illness severity parameters, as well as four types of age-specific contact patterns at home, at work, at school, and at other locations (Davies, Klepac et al 2020), exploiting an existing global data set which includes Pakistan-specific 5-year age-group contact rate matrices (CRMs) for work contacts (W-CRMs), school contacts (S-CRMs), home contacts (H-CRMs), and other social contacts (O-CRMs) (Prem, Cook & Jit 2017). Our transmission-dynamic epidemiological model framework has previously been applied to analysing NPI-related CRM-changes in the UK (Davies, Kucharski et al 2020), in three African settings including Mauritius, Nigeria and Niger (van Zandvoort et al 2020), and in Pakistan (Pearson et al 2020). However, in order to separately model province- and territory-specific epidemics, we exploited detailed population data from the WorldPop database (WorldPop 2020) to produce a bespoke set of baseline province- and territory-specific epidemic projections. In order to further improve projections, we adjusted the basic reproduction rate (R0) down from levels around R0=2.7, initially estimated for model set-up (Davies, Klepac et al 2020), to R0=1.7, in order to match Pakistan-specific R0 estimates (Uddin et al 2020, Ali, Imran & Khan 2020, Ullah & Khan 2020).^10^ In our policy simulations, business closure NPIs (BC-NPIs) are associated with changes in W-CRMs, school closure NPIs (SC-NPIs) are associated with changes in S-CRMs, home quarantine NPIs (HQ-NPIs) are associated with changes in H-CRMs, and group gathering NPIs (GG-NPIs) are associated with changes in O-CRMs.

In terms of our epidemiological model simulations, the CRM parametric assumptions, underlying our C19 baseline and three NPI policy scenarios 1-3, are presented in Table 2. The variation in scenario specifications involves (1) variation in reductions in W-CRM contact rates between −20% (Scenario 1) and −40% (Scenarios 2-3), and (2) variation in durations between one-off 10-week restrictions (Scenarios 1-2), and intermittent 1-month restrictions/2-month relaxations, of W-CRM, S-CRM, H-CRM and O-CRM contact rates. In terms of our macroeconomic model simulations of NPI interventions, needed to effectuate the reductions in contact-rates in policy scenarios 1-3, we assume (Table 3): (1) BC-NPI lockdowns affecting 20%/40% of non-essential businesses is required to achieve 20%/40% reduction in W-CRM contact-rates, (2) Nation-wide SC-NPI school closures are required to achieve 100% reductions in S-CRM contact-rates, (3) 10 day HQ-NPI home quarantines, combined with information campaigns (not modelled), are required to achieve 40% reductions in H-CRM contact-rates, and (4) nation-wide GG-NPI restrictions on group gatherings (not modelled) are required to achieve 20% reductions in O-CRM contact-rates. The durations of macroeconomic NPI interventions (Table 3) reflect the durations of the epidemiological CRM reductions over our 2020-21 time horizon (Table 2) and the timing of the Pakistani C19 epidemic outbreak in late-February 2020.

**Table 2.**
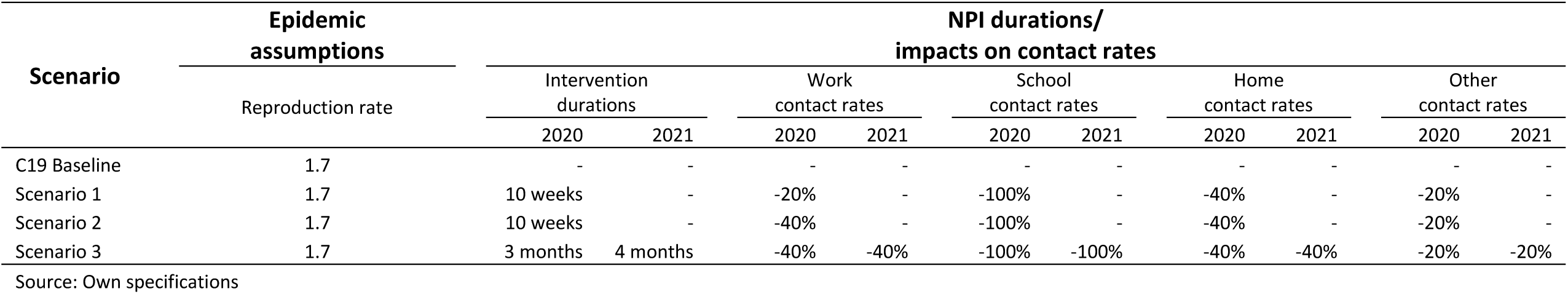
Epidemiological model scenario specifications (contact rate impacts by type of NPI)

**Table 3.**
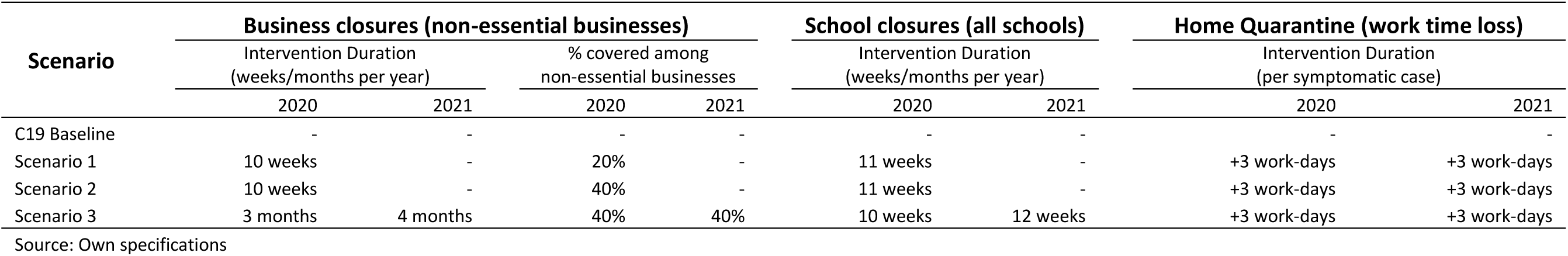
Macroeconomic model scenario specifications (by type of NPI)

| Scenario | Business closures (non-essential businesses) |  |  |  | School closures (all schools) |  | Home Quarantine (work time loss) |  |
| --- | --- | --- | --- | --- | --- | --- | --- | --- |
|  | Intervention Duration (weeks/months per year) |  | % covered among non-essential businesses |  | Intervention Duration (weeks/months per year) |  | Intervention Duration (per symptomatic case) |  |
|  | 2020 | 2021 | 2020 | 2021 | 2020 | 2021 | 2020 | 2021 |
| C19 Baseline | - | - | - | - | - | - | - | - |
| Scenario 1 | 10 weeks | - | 20% | - | 11 weeks | - | +3 work-days | +3 work-days |
| Scenario 2 | 10 weeks | - | 40% | - | 11 weeks | - | +3 work-days | +3 work-days |
| Scenario 3 | 3 months | 4 months | 40% | 40% | 10 weeks | 12 weeks | +3 work-days | +3 work-days |

Source: Own specifications

The epidemiological transmission-dynamic model provides outputs in terms of (1) non-hospitalized symptomatic cases; (2) severely ill hospitalized cases (not requiring ICU-treatment); (3) critically ill hospitalized cases (requiring ICU-treatment for part of their hospitalization duration); and (4) case fatalities. In order to derive macroeconomic health-related hospital cost- and labour supply-shocks, for our C19 baseline and three policy scenarios, we make the following parametric assumptions: (1) average cost of severely ill hospitalized patients is 33.32 USD/day; (2) average cost of critically ill hospitalized patients is 221.18 USD/day (Torres-Rueda et al 2020); (3) one-third of critical patient bed-days are treated in general ward while two-thirds are treated in ICU (ibid.), implying that the average cost of an ICU bed-day is 315.11 USD/day; (4) body bag cost for case fatalities is 64.52 USD/case fatality (ibid.); (5) non-hospitalized symptomatic cases have an average sickness duration of 10 weekdays/7 workdays; (6) severely ill and employed patients have a total illness duration of 15 weekdays/11 workdays; (7) critically ill and employed patients have a total illness duration of 17 weekdays/13 workdays; (8) employed case fatalities lose on average 105+250=355 workdays during 2020-21 if they die in 2020, and on average 125 workdays if they die in 2021. These eight parametric assumptions, together with age-specific labour force participation rates (PBS 2018), allow us to derive all health-related costs and labour force impacts, including (1) two separate decompositions of hospital admission costs (severely ill vs. critically ill hospitalization costs/non-ICU vs. ICU bed-day costs), (2) case fatality body bag costs, and (3) labour supply losses from employed non-hospitalized symptomatic cases, employed hospitalized severely and critically ill patients, and employed case fatalities.

The NPI interventions, needed to effectuate reductions in CRM contact rates, are quantified in the following way: (1) BC-NPI business closures for non-essential businesses are implemented by fixed proportional sector-specific factor demand reductions; (2) SC-NPI school closures’ impacts on parents’ labour force participation are assessed by dividing the number of province-specific school-age children (ages 5-14) by province-specific population-weighted average fertility rates (UN 2020), and multiplying by ‘fertility age’-weighted female labour force participation rates (PBS 2018), thereby assuming that one parent per average-sized family will be devoting their time to home schooling, and assuming that that parent will be the mother – an important conservative assumption given the significant gender-imbalance in Pakistani labour force participation rates (ibid.); (3) HQ-NPI home quarantine restrictions are assumed to be 14 days/10 workdays per non-hospitalized symptomatic case, thereby adding 3 additional workdays lost beyond our assumed C19 baseline 7 workday absenteeism per non-hospitalized symptomatic case.

## Results

The GDP impacts of our C19 baseline and three NPI policy scenarios 1-3 are tabulated in Tables 4-5, with aggregate impacts for 2020-21 tabulated in Table 4, and disaggregated impacts for 2020 and 2021 tabulated, separately, in Table 5. The aggregate results indicate that an unmitigated Pakistan C19 epidemic, by itself, would have led to a 0.12% reduction in Pakistani GDP (−721mn USD), and a total of 0.65mn critically ill and 1.52mn severely ill C19 patients; and 405,000 Pakistani citizens would have lost their lives to the disease during 2020-21.

**Table 4.**
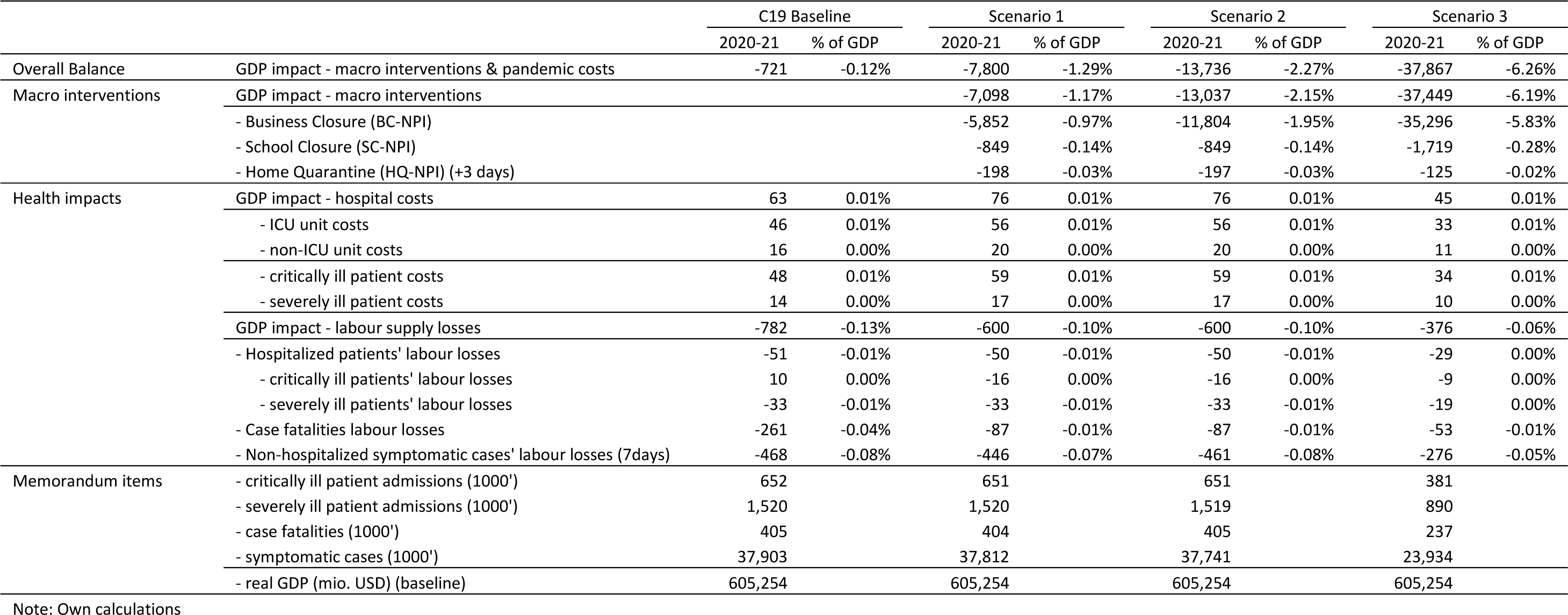
Macro-intervention and pandemic costs (GDP impacts for 2020-21; mio. USD) & trade-offs with clinical outcomes.

**Table 5.** Macro-intervention and pandemic costs (GDP impacts for 2020 and 2021; mio. USD) & trade-offs with clinical outcomes.

|  |  | C19 Baseline |  | Scenario 1 |  | Scenario 2 |  | Scenario 3 |  |
| --- | --- | --- | --- | --- | --- | --- | --- | --- | --- |
|  |  | 2020 | 2021 | 2020 | 2021 | 2020 | 2021 | 2020 | 2021 |
| Overall Balance | GDP impact - macro interventions & pandemic costs | -503 | -218 | -6,591 | -1,209 | -12,191 | -1,545 | -15,505 | -22,362 |
| Macro interventions | GDP impact - macro interventions |  |  | -6,566 | -532 | -12,172 | -865 | -15,483 | -21,966 |
|  | - Business Closure (BC-NPI) |  |  | -5,550 | -302 | -11,170 | -633 | -14,578 | -20,718 |
|  | - School Closure (SC-NPI) |  |  | -837 | -12 | -837 | -12 | -761 | -958 |
|  | - Home Quarantine (HQ-NPI) (+3 days) |  |  | -8 | -189 | -6 | -191 | -7 | -118 |
| Hospital Costs | GDP impact - hospital costs | 57 | 6 | 1 | 75 | 1 | 75 | 1 | 43 |
|  | - ICU unit costs | 41 | 5 | 1 | 55 | 1 | 55 | 1 | 32 |
|  | - non-ICU unit costs | 15 | 2 | 0 | 19 | 0 | 19 | 0 | 11 |
|  | - critically ill patient costs | 44 | 5 | 1 | 58 | 1 | 58 | 1 | 33 |
|  | - severely ill patient costs | 12 | 1 | 0 | 16 | 0 | 16 | 0 | 9 |
| Labour Supply losses | GDP impact - labour supply losses | -559 | -223 | -21 | -579 | -16 | -584 | -19 | -357 |
|  | - Hospitalized patients' labour losses | -46 | -5 | -1 | -49 | -1 | -49 | -1 | -28 |
|  | - critically ill patients' labour losses | 8 | 2 | 0 | -16 | 0 | -16 | 0 | -9 |
|  | - severely ill patients' labour losses | -30 | -3 | -1 | -32 | 0 | -32 | -1 | -18 |
|  | - Case fatalities labour losses | -74 | -188 | -1 | -86 | -1 | -86 | -1 | -52 |
|  | - Non-hospitalized symptomatic cases' labour losses (7days) | -439 | -29 | -5 | -441 | -14 | -446 | -2 | -275 |
| Memorandum items | - critically ill patient admissions (1000') | 582.9 | 68.7 | 12.0 | 638.6 | 8.9 | 641.7 | 13.3 | 367.6 |
|  | - severely ill patient admissions (1000') | 1366.2 | 154.2 | 29.4 | 1490.6 | 21.8 | 1497.2 | 32.1 | 858.0 |
|  | - case fatalities (1000') | 345.3 | 60.0 | 4.5 | 399.9 | 3.3 | 401.7 | 6.7 | 230.4 |
|  | - symptomatic cases (1000') | 35,902.0 | 2,000.7 | 1,745.4 | 36,066.3 | 1,274.6 | 36,466.3 | 1,410.6 | 22,523.6 |
|  | - real GDP (mio. USD) (baseline) | 298,440 | 306,814 | 298,440 | 306,814 | 298,440 | 306,814 | 298,440 | 306,814 |

Note: Own calculations

Our one-off 10-week lockdown scenarios 1-2 would only have postponed (Table 5), but not lessened (Table 4), the aggregate disease burden, within our two year time horizon, and the cost to society would have been 10-20 times higher than the unmitigated baseline. Hence, while both of the NPI policy scenarios 1-2, similar to the unmitigated baseline, would have led to 0.65mn critically ill/1.52mn severely ill/404-405,000 case fatalities during 2020-21, their cost to Pakistani society would have increased by an order of magnitude (or more) to respectively 1.29% of GDP (7.8bn USD) for Scenario 1, and 2.27% (13.7bn USD) for Scenario 2. Based on these results, it is evident that a one-off time-limited 10-week lockdown would not have been effective in containing the Pakistani epidemic. In spite of their limited long-term success in containing transmission, infections, and deaths, these one-off NPI interventions might still have been cost-effective in terms of preserving public health. Under scenarios 1 and 2, the overwhelming majority of clinical presentations would have been delayed by around one year, the number of life years saved, from postponed case fatalities, would have been counted in the 100,000s, and both severe clinical and Long-COVID health repercussions would have been postponed for a year, bringing additional health benefits. For Scenario 1, for example, the NPI cost per saved life-year would have been <$20,000, indicating that this scenario, in spite of limited long-term containment of the epidemic, could still, possibly, have been cost-effective.

Turning to Scenario 3, the GDP cost of pursuing this intermittent lockdown scenario would have been >50 times higher than the unmitigated baseline, costing Pakistani society a total of 37.9bn USD (6.26% of GDP) or 37.1bn USD (6.14% of GDP) more than the baseline during 2020-21 (Table 4). However, the recurring nature of the strategy also means that the aggregate long-term disease burden would have been reduced at the end of our two year time horizon, including reductions in critically ill C19 patients from 0.65mn to 0.38mn (−42%), severely ill C19 patients from 1.52mn to 0.89mn (−43%), and case fatalities from 405,000 citizens to 237,000 citizens (−42%). This suggests that our intermittent scenario, although costly both in pecuniary terms and in lives lost, could be an, at least partially, effective strategy for long-term containment and control of the Pakistani C19 epidemic. That being said, the implied reduction in case fatalities, over our two-year horizon, ‘only’ amounts to 41.5%, and the number of life-years saved are ‘only’ likely to be in the order of 500,000, indicating a cost per life saved of around $75,000. Hence, the number of lives saved by the hard broad-based intermittent lockdown strategy may not be sufficiently large, for it to be considered as cost-effective compared with (less costly) healthcare interventions.

Taking a longer-term perspective, and factoring-in the future roll-out of vaccination programmes, this conclusion may, however, change quite dramatically. If vaccine-rollout was to be completed by end-of-2021, the postponement of infections and deaths may transition from being temporary to being permanent, and the halving of case fatalities in Scenario 3, compared to Scenario 1, could result, not just in >200,000 life-years saved during 2020-21, but possibly millions of life-years saved over the coming decade. Thus, subject to confounding by virus mutations and their resulting requirement for vaccine re-development and additional vaccine roll-out programmes, our intermittent lockdown scenario 3 might be considered to be cost-effective and preferable to our one-off 10-week lockdown scenario 1 (even if vaccination programme costs end up running into billions USD).

Looking at scenario cost dynamics between 2020 and 2021, (Table 5), our results indicate that an unmitigated epidemic would incur overall GDP costs of 0.5bn USD in 2020 and 0.2bn USD in 2021 (70%/30%). In contrast the costs incurred for other scenarios would be 6.6/1.2bn USD (85%/15%) for scenario 1, 12.2/1.5bn USD (89%/11%) for scenario 2 and 15.5/22.4bn USD (45%/55%) for scenario 3. The BC-NPI costs in Table 5 (row three) highlight that these cost dynamics are driven by the timing of business lockdowns, indicating that employing a hard broad-based intermittent lockdown strategy, akin to scenario 3, may only be cost-effective over the very long-term (beyond our two-year time horizon), if efficacious vaccine roll-out is used to limit the future need for BC-NPIs.

Looking at the shorter perspective and comparing our epidemiologically simulated numbers of symptomatic cases and case fatalities for 2020 (Table 5), for Scenario 1 (1.75mn symptomatic cases & 4,500 fatalities), Scenario 2 (1.27mn symptomatic cases & 3,300 fatalities) and Scenario 3 (1.41mn symptomatic cases & 6,700 fatalities), with observed cumulative Pakistani outcomes on 31. December 2020 (482,506 confirmed cases & 10,176 fatalities) (GoP 2021), suggests that Pakistan’s smart lockdown strategy may not have fully accomplished the reduction in reproduction number and clinical outcomes that our hard broad-based lockdown scenarios could have accomplished. Hence, the number of case fatalities is around 50% higher than would have been accomplished with the intermittent 1-month lockdown/2-months opening-up scenario 3, and 2-3 times as high as what might have been accomplished by a full 10-week lockdown in early spring of 2020.

Looking at the individual cost components of our policy scenarios (Tables 4-5), NPI interventions make up 91-99% of total GDP costs, covering BC-NPIs (85%-95%), SC-NPIs (5-12%) and HQ-NPIs (0-3%). This confirms that BC-NPI business closures dominate GDP costs in the non-smart society-wide lockdown strategies, analysed here, and especially so in the intermittent lockdown scenario 3. Nonetheless, the results also show that HQ-NPI quarantine restrictions, and, especially, SC-NPI school closures, represent non-negligible GDP costs, and that combined non-BC-NPI GDP costs exceed unmitigated baseline costs by a factor of 1.5 (Scenarios 1-2) to 2.6 (Scenario 3).

Looking at the health-related GDP costs (Table 4), which include hospital costs and labour supply losses, it is interesting to note that increasing hospital costs provide a (small) economic stimulus (the induced shift in final demand composition, towards health services, seems to bring efficiency gains due to Pakistan being a second-best policy environment), while labour force losses, due to C19 illness and death, creates a larger drag on GDP. Overall, net health-related GDP costs for the uncontrolled baseline scenario amount to 721mn USD. Labour-related GDP losses dominate health-related costs, and they vary between 782mn USD (baseline), 600mn USD (Scenarios 1-2) and 376mn USD (scenario 3) (Table 4).

Tables 6-7 provide a breakdown of final demand drivers of GDP impacts. Looking at the GDP impacts of BC-NPI restrictions, they confirm that reduced real private consumption (RCP) (0.8-4.9% of real GDP) drives real GDP impacts, and accounts for around 84% of the total final demand reduction (not shown). Since RCP accounts for 86% of final demand along the counterfactual growth path (Table 6), RCP contributes proportionally to the final demand/real GDP reductions of BC-NPI interventions. Interestingly, the other domestic final demand components change disproportionally, including increases in real government consumption (RCG) (0.1-0.5% of real GDP) and relatively strong reductions in real investment (RINV) (0.2-1.4% of real GDP), while real import and real export impacts even out due to our fixed BoP external macro-closure (see appendix C). The systematic increases in RCG are conditioned by our government budget closure (see appendix C), i.e. government consumption being fixed at the counterfactual growth path and only varying with scenario-specific changes in hospital costs. Hence, since declining employment opportunities in non-essential sectors are likely to drive down wages, and, by implication, reduce costs of essential public services (i.e. the government price deflator), government service delivery opportunities increase within a fixed government budget environment.

**Table 6.** Macro-interventions breakdown (GDP decomposition impacts for 2020-21; mio. USD)

|  |  | Scenario 1 |  | Scenario 2 |  | Scenario 3 |  |
| --- | --- | --- | --- | --- | --- | --- | --- |
|  |  | 2020-21 | % of GDP | 2020-21 | % of GDP | 2020-21 | % of GDP |
| Macro interventions |  | -7,098 | -1.17% | -13,037 | -2.15% | -37,449 | -6.19% |
| - Business Lockdown | Δreal GDP | -5,852 | -0.97% | -11,804 | -1.95% | -35,296 | -5.83% |
|  | Δreal Private Consumption | -4,938 | -0.82% | -9,923 | -1.64% | -29,606 | -4.89% |
|  | Δreal Government Consumption | 490 | 0.08% | 928 | 0.15% | 2,874 | 0.47% |
|  | Δreal Investment | -1,404 | -0.23% | -2,809 | -0.46% | -8,564 | -1.41% |
|  | Δreal Export | -1,365 | -0.23% | -2,716 | -0.45% | -8,131 | -1.34% |
|  | Δreal Import | -1,365 | -0.23% | -2,716 | -0.45% | -8,131 | -1.34% |
| - School Closure | Δreal GDP | -849 | -0.14% | -849 | -0.14% | -1,719 | -0.28% |
|  | Δreal Private Consumption | -743 | -0.12% | -743 | -0.12% | -1,504 | -0.25% |
|  | Δreal Government Consumption | -25 | 0.00% | -25 | 0.00% | -49 | -0.01% |
|  | Δreal Investment | -82 | -0.01% | -82 | -0.01% | -165 | -0.03% |
|  | Δreal Export | -188 | -0.03% | -188 | -0.03% | -383 | -0.06% |
|  | Δreal Import | -188 | -0.03% | -188 | -0.03% | -383 | -0.06% |
| - Quarantine (10 days) | Δreal GDP | -198 | -0.03% | -197 | -0.03% | -125 | -0.02% |
|  | Δreal Private Consumption | -173 | -0.03% | -173 | -0.03% | -109 | -0.02% |
|  | Δreal Government Consumption | -6 | 0.00% | -6 | 0.00% | -4 | 0.00% |
|  | Δreal Investment | -19 | 0.00% | -19 | 0.00% | -12 | 0.00% |
|  | Δreal Export | -44 | -0.01% | -44 | -0.01% | -28 | 0.00% |
|  | Δreal Import | -44 | -0.01% | -44 | -0.01% | -28 | 0.00% |
| Memorandum items: | real GDP | 605,254 | % of GDP | 605,254 | % of GDP | 605,254 | % of GDP |
|  | Δreal Private Consumption | 518,393 | 85.6% | 518,393 | 85.6% | 518,393 | 85.6% |
|  | Δreal Government Consumption | 68,284 | 11.3% | 68,284 | 11.3% | 68,284 | 11.3% |
|  | Δreal Investment | 73,687 | 12.2% | 73,687 | 12.2% | 73,687 | 12.2% |
|  | Δreal Export | 83,941 | 13.9% | 83,941 | 13.9% | 83,941 | 13.9% |
|  | Δreal Import | 139,051 | -23.0% | 139,051 | -23.0% | 139,051 | -23.0% |

Note: own calculations

**Table 7.**
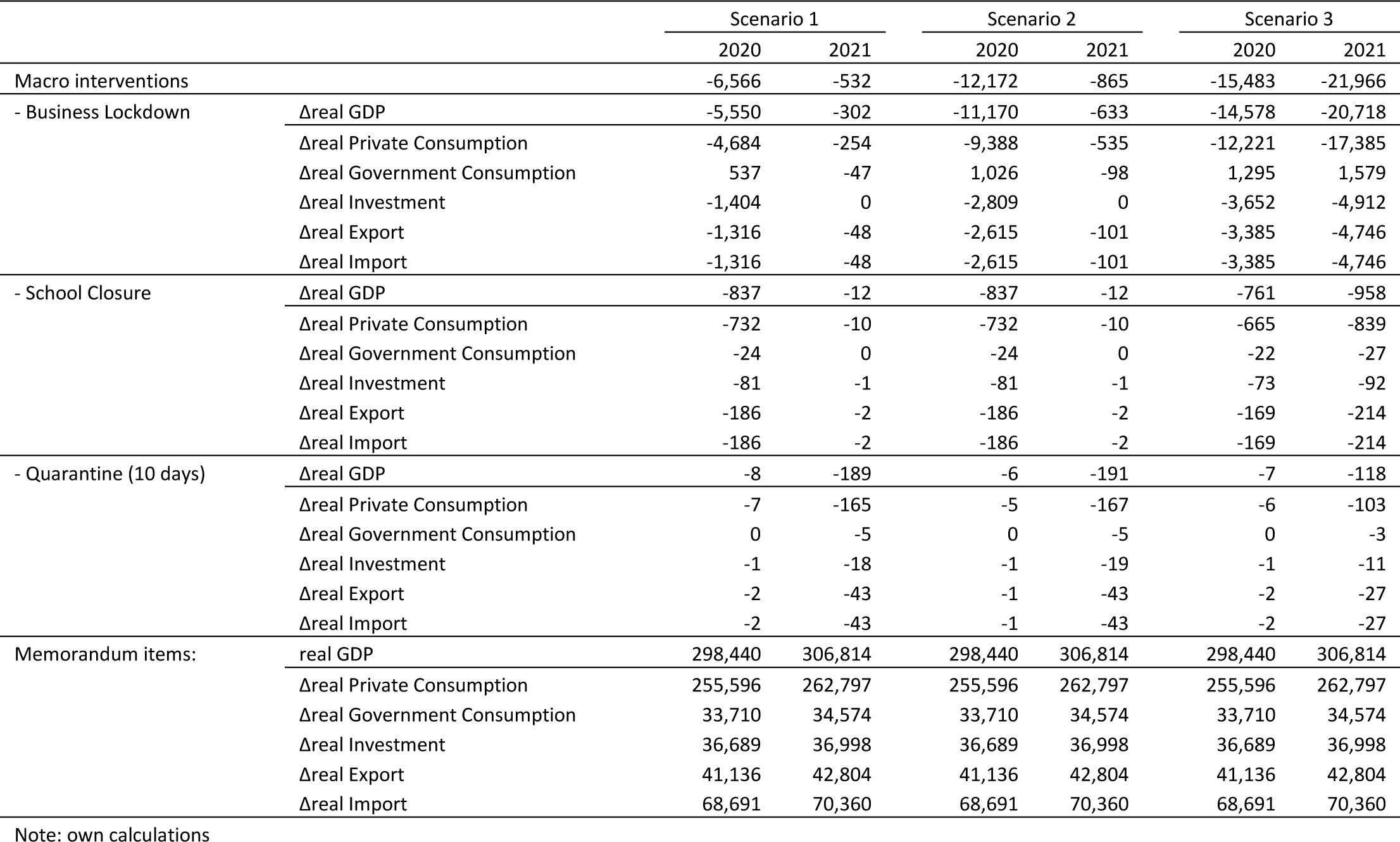
Macro-interventions breakdown (GDP decomposition impacts for 2020 and 2021; mio. USD)

In contrast, declines in RINV are relatively large, accounting for 24% of the total final demand reduction (not shown). Since RINV accounts for 12% of final demand along the counterfactual growth path (Table 6), it contributes disproportionally to the final demand (and real GDP) reductions of BC-NPI interventions. The supply-side driven BC-NPI sector lockdowns, including non-essential investment goods sectors, drives up investment prices, and this drives down real investment demand (beyond the reduction in private savings). The implications are three-fold: (1) 2020 real GDP declines due to reduced same-period investment demand, (2) 2021 GDP declines due to reduced 2020 capital accumulation, and reduced 2021 investment demand, and (3) longer-term GDP (beyond our two-year time horizon) declines are expected due to the reduced 2020-21 capital accumulation.

Turning to the SC-NPI and HQ-NPI restrictions, they do not involve BC-NPI-style sector-specific supply-side restrictions. While it has already been established that the labour force impacts of these restrictions are notable, reducing real GDP by more than the unmitigated epidemic, itself, these restrictions do not affect economic sectors asymmetrically. By implication, all final demand components are reduced proportionally (more or less), including RCP and RINV accounting for respectively 87-88% and 10% of final demand reductions. The smaller relative real investment impacts imply smaller long-term growth impacts, compared to the BC-NPI restrictions, suggesting that the SC-NPI and HQ-NPI restrictions, in combination with Pakistan’s preferred smart business lockdown-measures, may be a preferred strategy.

A regional breakdown of health-related GDP impacts across the four main regions of Pakistan (Tables 8-9) shows that hospital and patient-related real GDP losses are shared across regions in proportion to regional population size and clinical outcome numbers. If the Pakistan epidemic had not been controlled, our baseline simulation indicates that Punjab, being the largest region, would have expanded hospital activity the most, and, by implication, also expanded real GDP the most (36mn USD), followed by Sindh (16mn USD), Balochistan (3mn USD), and KPK (3mn USD), but these economic ‘benefits’ would have been outweighed by stronger regional labour force-related GDP losses, including Punjab (495mn USD), Sindh (188mn USD), Balochistan (37mn USD) and KPK (22mn USD). As already mentioned, aggregate labour force GDP losses decline from 782mn USD (unmitigated baseline) to 600mn USD in scenarios 1-2 (23% reduction), and to 376mn USD in scenario 3 (52% reduction), and our province-level results confirm that these reductions (except for minor non-linearities) would be shared, almost proportionally, across the four provinces. This demonstrates the potential for NPI policy scenarios to simultaneously lower health burdens and health-related economic burdens, and to do it in an equitable way, across the four provinces of Pakistan. Obviously, this conclusion has to be tempered, as discussed at length, by the large GDP costs which would result from implementing the hard FL-style lockdown strategies, analysed here.

**Table 8.** Health impact breakdown across regions (GDP impacts for 2020-21; mio. USD)

|  |  | C19 Baseline |  | Scenario 1 |  | Scenario 2 |  | Scenario 3 |  |
| --- | --- | --- | --- | --- | --- | --- | --- | --- | --- |
|  |  | 2020-21 | % of GDP | 2020-21 | % of GDP | 2020-21 | % of GDP | 2020-21 | % of GDP |
| Health impacts | ΔGDP - labour Supply losses | -782 | -0.13% | -600 | -0.10% | -600 | -0.10% | -376 | -0.06% |
|  | - Household 1 - KPK | -22 | 0.00% | -17 | 0.00% | -17 | 0.00% | -11 | 0.00% |
|  | - Household 2 - PUNJAB | -495 | -0.08% | -383 | -0.06% | -382 | -0.06% | -239 | -0.04% |
|  | - Household 3 - SINDH | -188 | -0.03% | -173 | -0.03% | -172 | -0.03% | -107 | -0.02% |
|  | - Household 4 - BALOCHISTAN | -37 | -0.01% | -28 | 0.00% | -28 | 0.00% | -18 | 0.00% |
|  | ΔGDP - hospital Costs | 63 | 0.01% | 76 | 0.01% | 76 | 0.01% | 45 | 0.01% |
|  | - Household 1 - KPK | 3 | 0.00% | 3 | 0.00% | 3 | 0.00% | 2 | 0.00% |
|  | - Household 2 - PUNJAB | 42 | 0.01% | 50 | 0.01% | 50 | 0.01% | 29 | 0.00% |
|  | - Household 3 - SINDH | 16 | 0.00% | 20 | 0.00% | 20 | 0.00% | 12 | 0.00% |
|  | - Household 4 - BALOCHISTAN | 3 | 0.00% | 3 | 0.00% | 3 | 0.00% | 2 | 0.00% |
| Memorandum item: | real GDP | 605,254 |  | 605,254 |  | 605,254 |  | 605,254 |  |

Note: own calculations

**Table 9.** Health impact breakdown across regions (GDP impacts for 2020 and 2021; mio. USD)

|  |  | C19 Baseline |  | Scenario 1 |  | Scenario 2 |  | Scenario 3 |  |
| --- | --- | --- | --- | --- | --- | --- | --- | --- | --- |
|  |  | 2020 | 2021 | 2020 | 2021 | 2020 | 2021 | 2020 | 2021 |
| Health impacts | ΔGDP - labour Supply losses | -559 | -223 | -21 | -579 | -16 | -584 | -19 | -357 |
|  | - Household 1 - KPK | -16 | -6 | -3 | -14 | -2 | -14 | -3 | -8 |
|  | - Household 2 - PUNJAB | -350 | -145 | -6 | -376 | -4 | -378 | -6 | -233 |
|  | - Household 3 - SINDH | -125 | -63 | -7 | -166 | -5 | -167 | -6 | -101 |
|  | - Household 4 - BALOCHISTAN | -27 | -10 | -5 | -23 | -4 | -24 | -4 | -14 |
|  | ΔGDP - hospital Costs | 57 | 6 | 1 | 75 | 1 | 75 | 1 | 43 |
|  | - Household 1 - KPK | 3 | 0 | 0 | 3 | 0 | 3 | 0 | 2 |
|  | - Household 2 - PUNJAB | 36 | 6 | 0 | 50 | 0 | 50 | 0 | 29 |
|  | - Household 3 - SINDH | 16 | 1 | 0 | 20 | 0 | 20 | 0 | 11 |
|  | - Household 4 - BALOCHISTAN | 3 | 0 | 0 | 3 | 0 | 3 | 0 | 2 |
| memorandum item: | real GDP | 298,440 | 306,814 | 298,440 | 306,814 | 298,440 | 306,814 | 298,440 | 306,814 |

Note: own calculations

## Conclusion and Discussion

Our simulations suggest that an uncontrolled Pakistan C19 epidemic, by itself, would have led to a 0.12% reduction in Pakistani GDP (−721mn USD), and a total of 0.65mn critically ill and 1.52mn severely ill C19 patients during 2020-21, while 405,000 Pakistani citizens would have lost their lives to the disease over the same period. Since the majority of case fatalities and symptomatic cases, respectively 345,000 and 35.9mn, would have occurred in 2020, the case fatality and confirmed case numbers, actually achieved by 31. December 2020 (10,176 fatalities & 482,506 confirmed cases) (GoP 2021) and by 14. October 2021 (28,201 fatalities and 1,261,685 confirmed cases) (Ritchie 2021), represents an outcome which is much better than feared by Pakistani health officials during Spring 2020 when the national C19 epidemic took off.

When compared to our NPI policy scenarios, it may be surmised that, while Pakistan’s switch from forced broad-based lockdowns during 15-30. March 2020, to a less constrained partial/smart lockdown strategy from 31. March 2020, is likely to have lowered the macroeconomic costs of C19 control, it is equally likely to have been less favourable in terms of health outcomes. Both our forced one-off and intermittent strategies would have postponed the C19 epidemic until 2021, lowering symptomatic case numbers and case fatalities, during 2020, to respectively 1.27-1.74mn cases & 3,300-4,500 fatalities (scenarios 1-2), and to 1.41mn cases & 6,700 fatalities (scenario 3). Not considering the great uncertainty during the initial months of the C19 pandemic, the Pakistani authorities could therefore, potentially, have further lowered clinical outcomes and bought more time for the (at the time uncertain) vaccine development and subsequent vaccine roll-out. However, the potential reductions in case fatalities, ranging from 50% in intermittent scenario 3, to a two-fold/three-fold reduction in the 10-week forced lockdown scenarios 1-2, would have come at an NPI-related 2020 GDP cost to society amounting to between 6.6-15.5bn USD (2.2%-5.2% of 2020 GDP).

While the 2020 GDP cost of Pakistan’s smart lockdown strategy remains unclear, Pakistan’s real GDP growth rates had been declining, over past (fiscal) years, from 5.2% and 5.5% during 2016-17 and 2017-18, to 1.9% during 2018-19 (PBS 2021). Based on the trend decline in real GDP over previous years, and the downward revision of the 2018-19 growth rate to 1.0% and the subsequent estimate of 0.5% GDP growth during 2019-20 (WB 2021) combined with relatively favourable World Economic Outlook projections of 1.5% growth during 2020-21 published in January 2021 (IMF 2021), it would seem that Pakistan, during 2020, has managed to control their C19 epidemic at a relatively low GDP cost to Pakistani society. If the Pakistani authorities had adopted hard lockdown strategies, akin to our simulated policy scenarios, they could therefore have reduced clinical outcomes but only at the expense of much higher GDP costs, a large part of which would likely have affected low-income households, something which the Pakistani government has made great efforts to avoid, for example, via their Ehsaas social protection programme roll-out in early 2020. In general, the Pakistani national and provincial authorities seem to have taken a balanced approach to C19 epidemic control, during 2020, via management of contact numbers at Pakistani schools and within Pakistani households, to such an extent that it has allowed business closures to be limited to selected sectors, and this balanced approach, with a focus on at most 1-day-a-week or 2-day-a-week business lockdowns, limited to hotspot areas, and coordinated across provinces and territories by the NCOC National Command Operation Center, was successfully continued throughout the alpha and delta variant corona-waves of 2021.

Going forward, our results suggest that the national Pakistani C19 epidemic is likely to affect all parts of Pakistani society, perhaps with a bias towards urban areas with high population densities, but otherwise fairly equally across provinces. Furthermore, our results suggest that hard broad-based long-duration lockdowns are likely to be a costly approach to reduced human activity and C19 epidemic control. Given that the smart lockdown approach appears to have been successful during 2020-21, it would probably serve Pakistan well to continue the use of limited business closures, possibly complemented by more wide-ranging interventions to control contact numbers at schools, in Pakistani homes (through information campaigns), and in other locations, in order to ensure that interventions remain effective, but also keep the GDP cost of their lockdown strategy manageable, especially for poorer informal sector households. Resumption of the successful Ehsass transfer program could also be considered, but this would, probably, only be warranted if the now widespread transmission of the B117 strain were to further accelerate and turn into nation-wide exponential growth, with costly nation-wide broad-based long-duration lockdowns to follow.

In terms of impacts on the composition of final demand, our simulated NPI strategies were shown to have lop-sided impacts on real investment. Hence, within our policy scenarios, unchanged, or slightly increasing, real government consumption crowded-out real investment within a shrinking economy. This is important since declining real investment demand affects both current and future income generation via capital accumulation. Since business closure restrictions were seen to have a particularly lopsided impact on investment demand, a strategy composed of school closure and home quarantine restrictions, in combination with Pakistan’s preferred smart business lockdown-measures, may be a preferred strategy, both from a short-term informal worker economy-perspective, and from a longer-term formal economy growth perspective. Regardless of the NPI strategy, once effective epidemic control has been established, there will be a need, not only for short-term stabilization of final demand, for example, via increased public investment demand for productive infrastructure projects, but also for stimulating private investment demand e.g. via fiscal policy measures to improve business returns, and for attracting foreign investment e.g. via measures to lower uncertainty about future market conditions.

The main lesson, from our policy scenarios, is that there is a clear trade-off between Covid-19 epidemic control and macroeconomic outcomes in Pakistan, and, potentially, in other LMICs, where access to pharmaceutical interventions is limited and where epidemic control is conditioned on non-pharmaceutical interventions. This trade-off is mainly conditioned by the extent to which human activity, measured by numbers of human contacts, can be controlled outside of workplaces. Hence, to the extent that a given population, at the behest of health authorities, accepts to limit their social contacts, at home and in other non-workplace settings, during peak transmission periods, this will lessen the need for broad-based business-closures, and allow for less costly ‘smart lockdown’ approaches with a focus on shorter and more geographically focussed reductions in non-business human contacts.

A corollary to the above is that subsequent pharmaceutical interventions, including the on-going roll-out of vaccines in Pakistan and abroad, is likely to have the highest value in societies where populations, for one reason or another, are not willing or able to reduce non-business contacts, for example refusing to abide by group gathering restrictions, during peak transmission periods. In these types of societies, the need for longer and more costly business-lockdowns are required, in order to avoid excessive pressures on health systems and excessive increases in excess mortality rates – and, by implication, the GDP value of vaccine roll-out, in these societies, is greater. This is not to say that the loss in quality of life, among populations with greater capacity for reducing social contacts, is not as great (or greater), and that these societies will not experience equally great (or greater) increases in quality of life from vaccine roll-outs, but the GDP value of vaccines is, nonetheless, likely to be greater in societies with less capacity for reducing social contacts.

A further corollary is that, while vaccine roll-outs may be prioritized higher, on economic grounds, in societies with less capacity for reducing social contacts, these societies are also likely to perform vaccine roll-out in a high-transmission setting, implying that, if national vaccine roll-out programs, for some reason, were to proceed slowly or be unable to reach heard immunity levels e.g. due to vaccine supply interruptions or vaccine scepticism, there would be an elevated risk of vaccine-specific mutations. By the same token, slow vaccine roll-out in populous countries, such as Pakistan, with seeming greater capacity for reducing social contacts, but with limited capacity for timely access to vaccines at scale, may also lead to long-term co-existence of pools of vaccinated and infectious individuals, and, thereby, to an elevated risk of vaccine-specific mutations and avoidable C19-related deaths. While the problem of dealing with avoidable deaths is, in principle, a domestic problem, virus mutations is a global problem, and the global community therefore has not only a moral responsibility, but a self-interest, in ensuring the timely provision of efficacious vaccines at scale to every country and territory around the world, including countries like Pakistan where the past year has seen nation-wide community-transmission of the highly infectious B117 alpha and B1617 delta variants, and where the smart lockdown strategies, which have worked so well during the first 1½ years of the Pakistan epidemic, may not be able to prevent long-term co-existence of large infectious and vaccinated population groups.

Several caveats apply to our analyses. While our economic assessments can be modified to encompass the consequences of changes to many types of social and economic NPI interventions, including business and school closures, it should be recognized that our approach disregards trade-offs in terms of important non-monetary impacts such as mental health for patients, relatives, health workers and the broader public, and (lack of) timely hospital treatment of non-C19 patients. Evidence is emerging regarding the potential longer-term Long-COVID post-C19 health sequelae (Huang et al 2021, Iqbal et al 2021, Ahmed, Patel et al 2020, Perrin et al 2020) and negative health system spillovers on other healthcare tasks during peak transmission periods, including treatment and screening for various cancer types (Skovlund et al 2021). Since these post-C19 manifestations and health system spillovers are still being studied, we did not account for them in our simulation analyses, but these twin areas do represent important areas of future research, in order to better understand longer-term post-C19 impacts on quality of life, and the resilience and preparedness of existing healthcare systems.

While full model integration has been achieved for macroeconomic modelling of other infectious diseases such as malaria (Jensen et al 2018, Smith et al 2020), knowledge gaps regarding socioeconomic feedback effects on epidemiological model parameters including human activity and contact matrices, currently bar full C19 epidemiological model integration. Furthermore, our approach to health-related GDP cost assessment, with its focus on labour force participation and hospital costs, is conservative in the sense that it excludes other potential transmission channels, for example, G-Cubed model channels such as increased equity risk premia, increased sector-specific (transport) costs of production, and changes in consumer preferences (McKibbin & Fernando 2020). That being said, the G-Cubed model simulations of 2-8% global C19 health-related GDP costs (ibid.) do not seem to have been borne out by reality in e.g. Pakistan. Going forward, the main methodological challenges will be to investigate the importance of economic feedback effects from, for example, household income and poverty levels to epidemiological model parameters, including age-stratified matrices for work, school, home, and other social contacts, but also to explore the validity of additional pathways for health-related GDP costs.

Although we do employ a validated reproduction number of 1.7, mirroring existing Pakistan-specific evidence, future research should look into the continuing transmission of variants of concern, including the B1617 delta variant, and their impacts on basic reproduction numbers. Our analyses could also possibly have been strengthened by explicit fitting of our epidemiological model to historical clinical trends. However, whilst this might have provided slightly different health burden and policy impact results, it would have been unlikely to substantially change our results and would also have been complicated by the need to disentangle smart lockdown impacts in order to derive a counterfactual model suitable for our simulations of uncontrolled and NPI-controlled epidemic scenarios. Since the purpose of this study is to present policy analyses of specific NPI scenarios, rather than forecasting of the future Pakistan C19 epidemic, we therefore decided to limit ourselves to application of our non-fitted epidemiological model framework. Similarly, due to limited knowledge of seasonal contact patterns, and complexities of modelling dynamic changes in contact matrices, most C19 epidemiological modelling applications (including our own) do not capture seasonality. As data becomes available, future applications could therefore also extend our analysis to capture policy-induced changes in timing and steepness of (winter) peaks and their implications for seasonal hospital capacity utilization, and better understand the optimal timing and duration of NPI policy interventions, in order to better support NPI epidemic control decision making.

In terms of future C19 control strategies, for Pakistan and other LMICs, the key issue involves vaccine roll-out in the context of potential delays in access to (efficacious) vaccine deliveries, and potential waning of vaccine efficacy either due to natural rates of immune response waning, or due to potential introduction or development of mutated C19 strains with increased resistance to vaccine-induced antibody responses. Due to the very real threats of slow vaccine roll-out and vaccine waning, health authorities in Pakistan and elsewhere should continue to maintain an epidemic preparedness infrastructure, which allows for rapid deployment of NPIs in the advent of future C19 resurgence and acceleration in transmission growth rates. While Pakistan has a good track-record of C19 epidemic control, so far (until early-November 2021), such a resurgence scenario is a very real possibility given the recent nation-wide community-transmission of the B1617 delta variant throughout Pakistan – and, given that vaccine roll-is still far from having achieved herd immunity, this pattern may well repeat itself over the coming years. Planning for such a scenario, including continued vigilance in the design of the vaccination roll-out, including continued focus on managing incentives for fellow citizens to take both the first and second jab, and potential future re-vaccination campaigns with intermittent smart lockdowns to control periods of C19 resurgence, should therefore be the focus of future C19 modelling efforts, especially in Pakistan and other LMICs, but also in HICs due to the unavoidable spread of mutated strains in a globalized world. The design of re-vaccination campaign strategies should also be informed by the experience of countries like Pakistan, where agile management of social contacts outside of the workplace, combined with sector-specific and geographically targeted smart lockdowns and effective central coordination, have so far managed to control their national C19 epidemics in a cost-effective way. That being said, Pakistan continues to face a reality where they have only recently managed to reduce country-wide community-transmission of the highly infectious B1617 variant, and, in spite of the successful move by national authorities to provide nation-wide lockdown guidelines, both provincial and national health authorities should consider re-examining existing control strategies, including a possible strengthening of focus on the management of non-business human activity in Pakistani society, and on continuously maintaining effective local pandemic preparedness systems.

## Data Availability

We will share the models upon request. The macroeconomic model contains data from the Global Trade Analysis Project (GTAP) which are proprietary, so cannot be shared along with the model.

https://doi.org/10.5281/zenodo.8411064

## Acknowledgements

This research was funded by a “Covid-19 Modelling for Pakistan” grant from the Foreign Commonwealth & Development Office, United Kingdom (Funder reference number 300796). The Foreign Commonwealth & Development Office had no role in the design, analysis or writing of this article. None of the authors have any conflict of interest of any type with respect to this manuscript.

## Appendix A. SAM accounts

**Table A.1.**
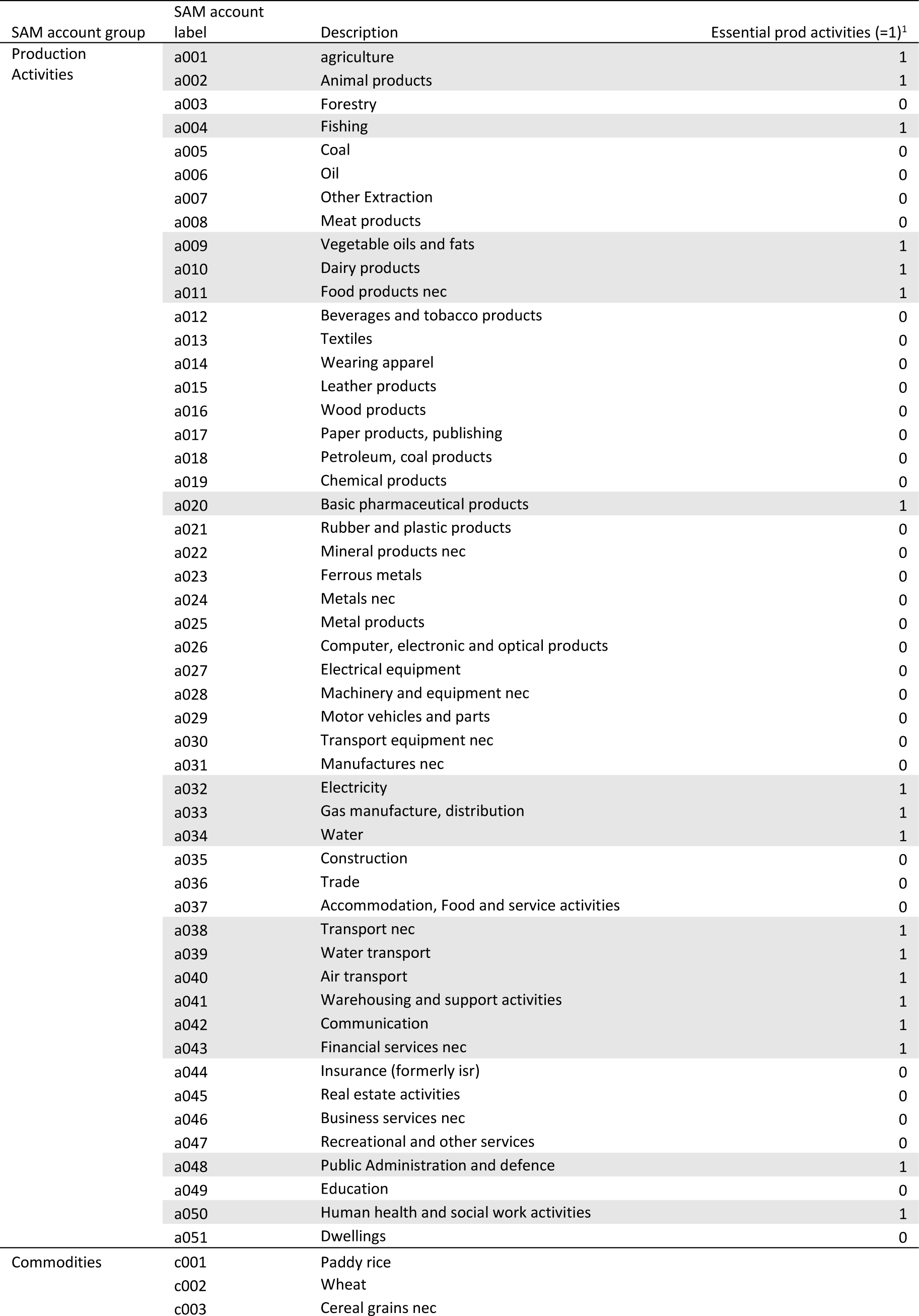
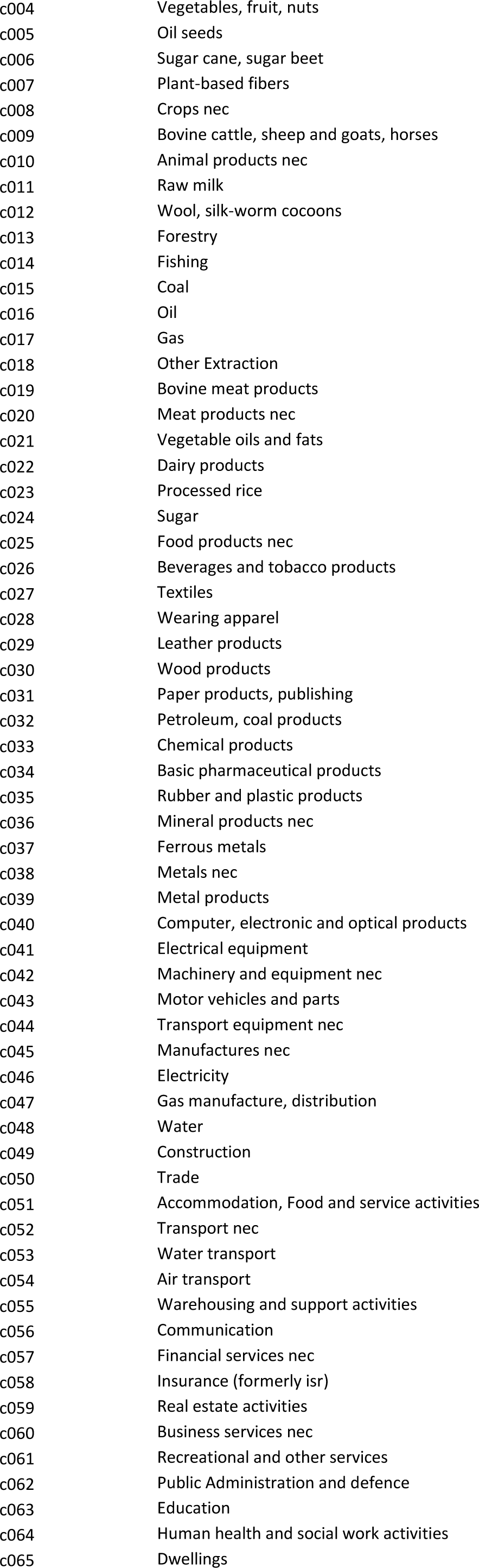
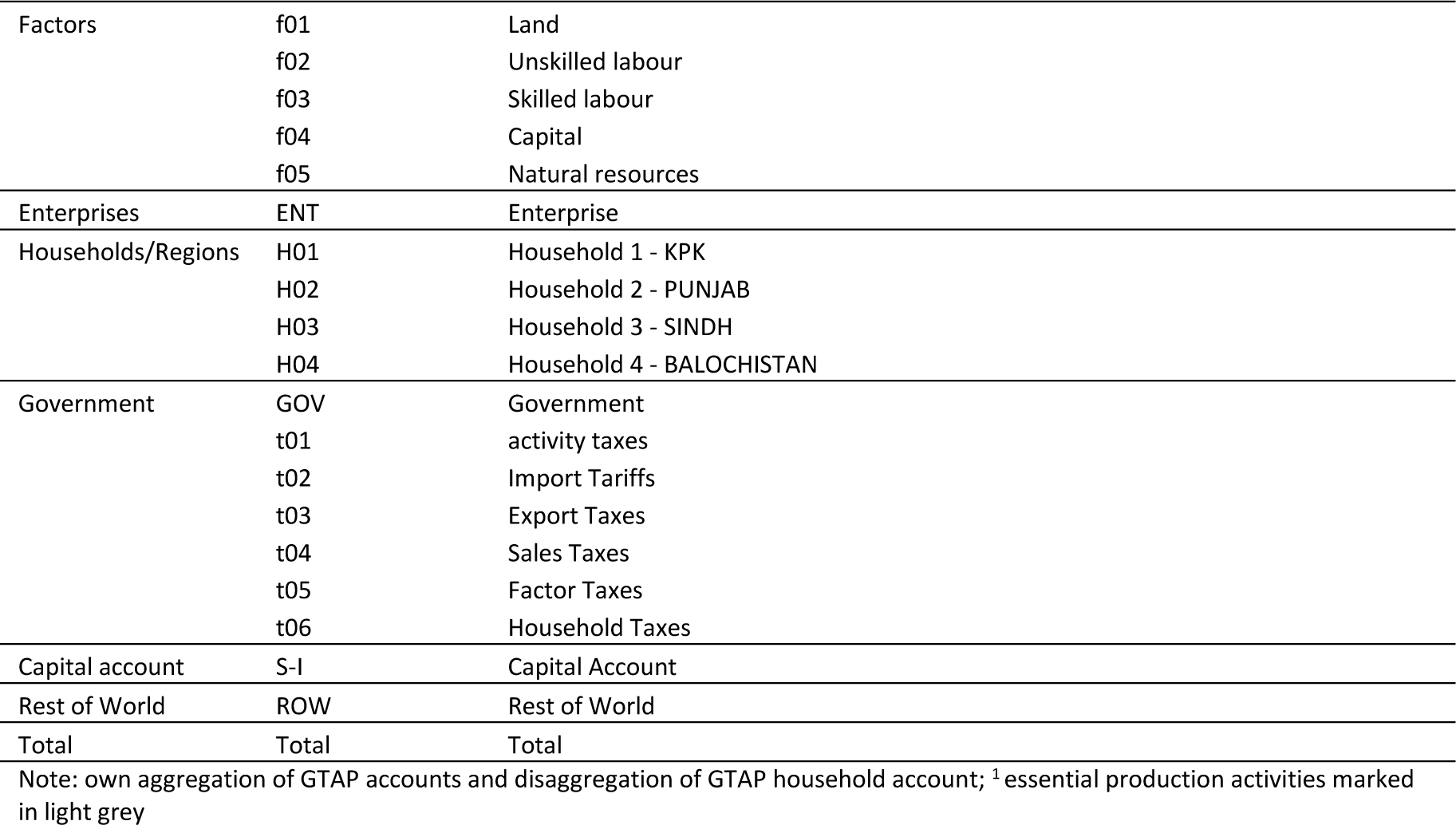
SAM accounts & essential production activities exempted from NPIs.

| SAM account group | SAM account label | Description | Essential prod activities (=1)<sup>1</sup> |
| --- | --- | --- | --- |
| Production Activities | a001 | agriculture | 1 |
|  | a002 | Animal products | 1 |
|  | a003 | Forestry | 0 |
|  | a004 | Fishing | 1 |
|  | a005 | Coal | 0 |
|  | a006 | Oil | 0 |
|  | a007 | Other Extraction | 0 |
|  | a008 | Meat products | 0 |
|  | a009 | Vegetable oils and fats | 1 |
|  | a010 | Dairy products | 1 |
|  | a011 | Food products nec | 1 |
|  | a012 | Beverages and tobacco products | 0 |
|  | a013 | Textiles | 0 |
|  | a014 | Wearing apparel | 0 |
|  | a015 | Leather products | 0 |
|  | a016 | Wood products | 0 |
|  | a017 | Paper products, publishing | 0 |
|  | a018 | Petroleum, coal products | 0 |
|  | a019 | Chemical products | 0 |
|  | a020 | Basic pharmaceutical products | 1 |
|  | a021 | Rubber and plastic products | 0 |
|  | a022 | Mineral products nec | 0 |
|  | a023 | Ferrous metals | 0 |
|  | a024 | Metals nec | 0 |
|  | a025 | Metal products | 0 |
|  | a026 | Computer, electronic and optical products | 0 |
|  | a027 | Electrical equipment | 0 |
|  | a028 | Machinery and equipment nec | 0 |
|  | a029 | Motor vehicles and parts | 0 |
|  | a030 | Transport equipment nec | 0 |
|  | a031 | Manufactures nec | 0 |
|  | a032 | Electricity | 1 |
|  | a033 | Gas manufacture, distribution | 1 |
|  | a034 | Water | 1 |
|  | a035 | Construction | 0 |
|  | a036 | Trade | 0 |
|  | a037 | Accommodation, Food and service activities | 0 |
|  | a038 | Transport nec | 1 |
|  | a039 | Water transport | 1 |
|  | a040 | Air transport | 1 |
|  | a041 | Warehousing and support activities | 1 |
|  | a042 | Communication | 1 |
|  | a043 | Financial services nec | 1 |
|  | a044 | Insurance (formerly isr) | 0 |
|  | a045 | Real estate activities | 0 |
|  | a046 | Business services nec | 0 |
|  | a047 | Recreational and other services | 0 |
|  | a048 | Public Administration and defence | 1 |
|  | a049 | Education | 0 |
|  | a050 | Human health and social work activities | 1 |
|  | a051 | Dwellings | 0 |
| Commodities | c001 | Paddy rice |  |
|  | c002 | Wheat |  |
|  | c003 | Cereal grains nec |  |

|  |  |
| --- | --- |
| c004 | Vegetables, fruit, nuts |
| c005 | Oil seeds |
| c006 | Sugar cane, sugar beet |
| c007 | Plant-based fibers |
| c008 | Crops nec |
| c009 | Bovine cattle, sheep and goats, horses |
| c010 | Animal products nec |
| c011 | Raw milk |
| c012 | Wool, silk-worm cocoons |
| c013 | Forestry |
| c014 | Fishing |
| c015 | Coal |
| c016 | Oil |
| c017 | Gas |
| c018 | Other Extraction |
| c019 | Bovine meat products |
| c020 | Meat products nec |
| c021 | Vegetable oils and fats |
| c022 | Dairy products |
| c023 | Processed rice |
| c024 | Sugar |
| c025 | Food products nec |
| c026 | Beverages and tobacco products |
| c027 | Textiles |
| c028 | Wearing apparel |
| c029 | Leather products |
| c030 | Wood products |
| c031 | Paper products, publishing |
| c032 | Petroleum, coal products |
| c033 | Chemical products |
| c034 | Basic pharmaceutical products |
| c035 | Rubber and plastic products |
| c036 | Mineral products nec |
| c037 | Ferrous metals |
| c038 | Metals nec |
| c039 | Metal products |
| c040 | Computer, electronic and optical products |
| c041 | Electrical equipment |
| c042 | Machinery and equipment nec |
| c043 | Motor vehicles and parts |
| c044 | Transport equipment nec |
| c045 | Manufactures nec |
| c046 | Electricity |
| c047 | Gas manufacture, distribution |
| c048 | Water |
| c049 | Construction |
| c050 | Trade |
| c051 | Accommodation, Food and service activities |
| c052 | Transport nec |
| c053 | Water transport |
| c054 | Air transport |
| c055 | Warehousing and support activities |
| c056 | Communication |
| c057 | Financial services nec |
| c058 | Insurance (formerly isr) |
| c059 | Real estate activities |
| c060 | Business services nec |
| c061 | Recreational and other services |
| c062 | Public Administration and defence |
| c063 | Education |
| c064 | Human health and social work activities |
| c065 | Dwellings |

|  |  |  |
| --- | --- | --- |
| Factors | f01 | Land |
|  | f02 | Unskilled labour |
|  | f03 | Skilled labour |
|  | f04 | Capital |
|  | f05 | Natural resources |
| Enterprises | ENT | Enterprise |
| Households/Regions | H01 | Household 1 - KPK |
|  | H02 | Household 2 - PUNJAB |
|  | H03 | Household 3 - SINDH |
|  | H04 | Household 4 - BALOCHISTAN |
| Government | GOV | Government |
|  | t01 | activity taxes |
|  | t02 | Import Tariffs |
|  | t03 | Export Taxes |
|  | t04 | Sales Taxes |
|  | t05 | Factor Taxes |
|  | t06 | Household Taxes |
| Capital account | S-I | Capital Account |
| Rest of World | ROW | Rest of World |
| Total | Total | Total |

Note: own aggregation of GTAP accounts and disaggregation of GTAP household account; <sup>1</sup> essential production activities marked in light grey

## Appendix B. Pakistan demographic and health situation

### Pakistan demographic and health trends

Pakistan experienced rapid population growth after independence in 1947. After moderate growth during the 1950s (1.55% p.a.), the demographic transition, with declining mortality rates, led to rapid population growth during the 1960s (4.3% p.a.), 1970s (3.6% p.a.) and into the 1980s (3.1% p.a.) Since the 1990s, average population growth rates have hovered around 2.3-2.6% p.a., due to fertility rates declining by an average −2.0% p.a. over the past three decades (Figure B.1) (GBD 2020). Recently accelerating declines in Pakistani age-specific fertility rates (ibid.) confirms that Pakistan can start to eye the end of her demographic transition. This can be verified, visually, by the broadening of the Pakistani population pyramid between 2010 and 2019 (Figure B.1). Pakistani population growth has also been accompanied by a changing demographic composition including a declining imbalance in the female-to-male gender ratio, and a population share of 65+ year olds which declined from 5.2% (1950) to 3.2% (2010) before increasing to 3.4% in 2019 (Figure B.1). Nonetheless, the Pakistani population remains fairly young with the 0-39 age group accounting for 81% of the population.

Pakistani disease burdens of major infectious diseases such as TB, HIV/AIDS, and malaria, although variable year-on-year, has been trending downwards over recent years (Figure B.2).

### National Pakistan disease background

Pakistan is a country which has been struggling with the twin challenges of a weak and underfunded health system and high population growth (see Figure B.1), and with twin CD and NCD burdens (see Figure B.2) (GBD 2020). However, the CD disease burden has been declining over the last couple of decades, and now amount to only around one-fifth of the total burden (ibid.)

Pakistan experienced rapid population growth after independence in 1947, including growth rates of 3-4% p.a. during the 1960s, 1970s, and 1980s. Since the 1990s, growth rates have hovered around 2.3-2.6% due to declining fertility rates (−2.0% p.a.), but, more recently, accelerating reductions in age-specific fertility rates have led to a renewed decline in population growth and a broadening of the Pakistani population pyramid. This confirms that Pakistan can start to eye the end of her demographic transition, and this is confirmed by declining imbalances in the female-to-male gender ratio, and a population share of 65+ year olds which, after declining from 5.2% (1950) to 3.2% (2010), has recently increased to 3.4% in 2019. Nonetheless, the Pakistani population remains fairly young with the 0-39 age group accounting for 81% of the population (GBD 2020).

In terms of demographics, a recent study found that the size of the 65+ age group is a significant factor in explaining the relative C19 caseloads in a cross-section of 182 countries (Nguiemkeu & Tadedjeu 2021). The relatively young population, and the fact that Pakistan has still not completed her demographic transition, may therefore provide a partial explanation for her relatively low C19 counts, so far, including 283 confirmed cases and 6.3 case fatalities per 100,000 population which compares well with 4000-9000 confirmed cases and 130-180 case fatalities per 100,000 population in other (select) populous countries (see Figure B.4). Other factors, identified by Nguiemkeu & Tadedjeu as significantly affecting relative C19 caseloads, include demographic and geographic variables such as population density (+), urban population share (+), and temperature (−), while they find socio-demographic variables such as GDP per capita and health expenditures to be insignificant (ibid.) The authors also note a caveat to their results, namely that “duration since start of epidemic” has a significantly positive secular impact on caseloads. Specifically, they demonstrate that Sub-Saharan Africa, in spite of having low initial caseloads, are experiencing a particularly high secular trend-increase in caseloads. While no attempt was made to undertake a separate analysis of South Asia, the identification of a high secular trend for Sub-Saharan Africa, with their relatively low confirmed caseloads and young populations, could be seen as a warning sign for Pakistan, that a young population and low initial caseloads, by themselves, may not shield her broader population in the longer term.

In terms of population density, the rapid post-independence population growth has increased the Pakistani population density from 59.8 to 275.3 people/km2 between 1950 and 2018, implying that Pakistan has moved from being 82^nd^ to being 41^st^ most densely populated country in the world, and suggesting that exposure to communicable disease has increased over time. At the same time, disease burdens of major infectious diseases such as TB, HIV/AIDS, and malaria, although variable year-on-year, has been trending downwards over recent years (see Figure B.2), indicating that improvements in sanitation and other preventive measures have, so far, allowed Pakistan to maintain control over the spread of communicable disease across her increasingly densely populated territory.

### Regional Pakistan disease background

After the 18^th^ constitutional amendment, passed in 2010, Pakistan consisted of eight regions, and after adoption of the 25^th^ constitutional amendment, passed in 2018 and leading to merger of the Federally Administered Tribal Areas (FATA) with Khyber Pakhtunkhwa (KP), Pakistan has consisted of seven administrative units including four provinces: Balochistan, Khyber Pakhtunkhwa (KPK), Punjab, and Sindh; and three territories including: Azad Jammu and Kashmir (AJK), Gilgit-Baltistan (GB), and Islamabad Capital Territory (ICT).

In terms of geography and climate, Pakistan’s territory can be broadly divided into three regions: the northern highlands (including KPK) with hot summers and cold winters (average winter temperatures <0 degrees Celsius), the Balochistan Plateau (including most of Balochistan but excluding the coastline) with hot summers and mild winters (average winter temperatures >0 degrees Celsius), and the fertile Indus River plains (including the populous Punjab/Sindh provinces) with hot summers and mild/very mild winters (average winter temperatures >0 degrees Celsius in Punjab and ICT, and >>0 degrees Celsius in Sindh).

While it has been suggested that climatic conditions, proxied by average temperatures, may condition the variation in caseloads between countries (Nguiemkeu & Tadedjeu 2021), the climatic variations between Pakistani provinces and territories do not seem to have created systematic variations in C19-epidemics between the regions of Pakistan. All four regions experienced accelerating transmission in late-October 2020 (see Figure B.3) suggesting that seasonality, rather than climate, were increasing transmission during the winter-months. However, in spite of the seasonally upward-sloping curves, relative levels of transmission and case fatalities, so far (by 11 March 2021), remain low by almost any standard.

The province- and territory-level data shows that all provinces and territories have, so far, managed to keep the C19-epidemic under relative control, perhaps with the exception of ICT which are showing the highest regional confirmed case count and case fatality rates amounting to respectively 1848 confirmed cases and 20.5 deaths per 100,000 population (per 11 March 2021). There are also slightly different epidemic patterns between, on the one hand, the four provinces (accounting for 95% of the Pakistani population) which quickly moved to gain control over the epidemic during June-July 2020, followed by a mild acceleration in transmission in late-October 2020 and mild constant growth during January-February 2021 (and, for Punjab, followed, in early-March, by strong re-acceleration in 7-day transmission growth rates from 2.5% (7-day period ending 28 February) to 4.3% (7-day period ending 11 March), and, on the other hand, the territories which, in the case of GB, only managed to break the curve in late-September 2020 (but, impressively, managed to maintain relative control throughout the winter-months), and, in the case of AJK and ICT, after seeing exponential increases in confirmed cases and case fatalities starting late-October 2020, they managed to stabilize, and, in the case of ICT decelerate, growth rates during January-February 2021 (but, similar to Punjab, a beginning re-acceleration of weekly transmission growth rates, in early-March 2021, from 2.0% (7-day period ending 28 February) to 4.2% (7-day period ending 11 March should be of concern for capital territory authorities).

Interestingly, the regional data demonstrates that ICT, apart from Balochistan, has the lowest observed Case Fatality Rates (CFR) per confirmed case (Table B.1). This can also be visually verified with reference to Figure B.3 (where primary and secondary vertical axis units have been normalized to match ICT numbers on 11 March 2021), by noting that regional case fatality-curves, for all other regions except Balochistan, has consistently dominated the confirmed case count-curves since June 2020.

The observed regional CFRs per confirmed case (per 11 March 2021) range from 1.04% in Balochistan and 1.11% in ICT, to 2.08% and 2.93% in the territories of GB and AJK, and to 2.85% and 3.12% in the KPK and Punjab provinces. Sindh province has an intermediate observed CFR of 1.71%. Benchmarking of observed regional CFRs against ICT, which has experienced both the highest (and strongest winter-acceleration in) confirmed case counts and case fatalities per 100,000 population, as well as the second-lowest observed CFR, suggests that (1) the highly urbanized nature of ICT is conducive for the spread of C19-virus, (2) that the advent of winter, with average minimum temperatures in the range of 3-5 degrees Celsius, is likely to have further increased transmission in ICT, and (3) the high concentration of health facilities in ICT may be keeping a lid on the CFR rate relative to other regions.

While the former hypothesis, linking urbanization with high transmission, is confirmed by other evidence discussed above (Nguiemkeu & Tadedjeu 2021), the latter hypothesis, linking concentration of tertiary health facilities with CFRs, may require nuancing in the current context. First of all, none of the regional epidemics have exceeded hospital capacity implying that differences in regional treatment capacity is unlikely to be responsible for the regional variation in CFRs. Another more likely explanation relates to differences in regional test capacity, which may lower numbers of confirmed cases in poorer regions such as Balochistan. It could also be hypothesized that less urbanized regions such as Balochistan, in spite of experiencing reduced transmission due to lower population density, should experience elevated CFRs due to greater average distances to tertiary health facilities. Such elevated CFRs may, however, not be observed due to more limited test capacity.

Interestingly, the two mountainous territories of GB and AJK, and the hilly province of KPK, have intermediate numbers of confirmed cases in the order of 230-458 per 100,000 and relatively high observed CFRs in the order of 2.08%-2.93%. Within this group of regions, the second-highest CFR (almost as high as AJK) and lowest confirmed case count-rate is observed in the hilly KPK province, and this suggests that most case fatalities with C19 infection are tested and confirmed in KPK, but also that mild cases may not be tested to the same extent as happens in ICT.

Most importantly, the two largest provinces, Punjab and Sindh, which accounts for a combined 77% of the Pakistani population, seems to have managed to control their province-level epidemics, so far, keeping confirmed case numbers down to 144-503 per 100,000 and maintain CFRs in the range 1.71-3.12%. The good overall Pakistani performance, so far, must be ascribed, partly, to the Punjabi and Sindh provincial governments, which seems to have been able to effectively contain their provincial epidemics, and, in spite of a relatively high CFR of 3.12% in Punjab, to keep observed case count-rates at low levels. While smart lockdown NPIs may have played a part in the impressive containment, so far, and while the Punjab and Sindh climate may have favoured out-door activities during at least the first part of the epidemic, until night-time temperatures started dropping below 20C towards end-of-October 2020, there is a need for province authorities to remain vigilant. Hence, in spite of the impressive containment, including the reduction of 7-day confirmed case count growth rates, which, after increasing from around 1% during the first week of November to between 4% (Punjab) and 7% (Sindh) during the first week of December, had been reduced to between 0.8-2.5% during the last week of February 2021, the Punjab and Sindh authorities would probably be well advised to continue to be vigilant because of the risk posed by accelerating transmission of the highly infectious B117 strain. Hence, in the case of Punjab, where B117 became dominant in several urban centres in early-March 2021, 7-day transmission growth rates re-accelerated from 2.5% (7-day period ending 28 February) to 4.3% (7-day period ending 11 March), and, due to its increased infectiousness and the lack of epidemic control instruments for containment, the strain is likely to become dominant throughout the rest of Punjab, Sindh, and the rest of Pakistan, during spring 2021.

### International disease context

Pakistan and South Asian (SA) countries, more broadly, have, so far, been relatively successful in controlling their C19-epidemics. This can be confirmed by international case count and case fatality comparisons vis-à-vis select countries, around the world, with large populations and with, in most cases, rampant and multi-wave epidemics (Figure B.4). By 11 March 2021, all SA countries had experienced relatively moderate-sized C19-epidemics as measured by case counts and case fatalities per 100,000 population. While India had recorded the fourth-highest number of confirmed case fatalities in the world (158,306), only surpassed by the US (530,821), Brazil (272,889), and Mexico (193,152), and vastly outnumbering Pakistan’s case fatality count (13,430) (Ritchie 2021), India’s population size meant that she had ‘only’ recorded 836 confirmed cases and 11.7 case fatalities per 100,000 population; and Pakistan’s record was even more ‘favourable’ with 283 confirmed cases and 6.3 case fatalities per 100,000 population. In a regional SA context, Pakistan ranked fifth and fourth, on 11 March 2021, in terms of case count and case fatalities per 100,000 population (Figure B.4), and the only SA country, with a uniformly better relative record at that time, was mountainous Bhutan, who had only recorded 115 cases and 0.1 case fatalities per 100,000 population (Ritchie 2021). Hence, both by international and regional SA standards, Pakistan’s C19 epidemic has, so far, been well-controlled, but the acceleration in community-transmission of B117, during early-March 2021 is, nonetheless, a cause for concern.

**Figure B.1.**
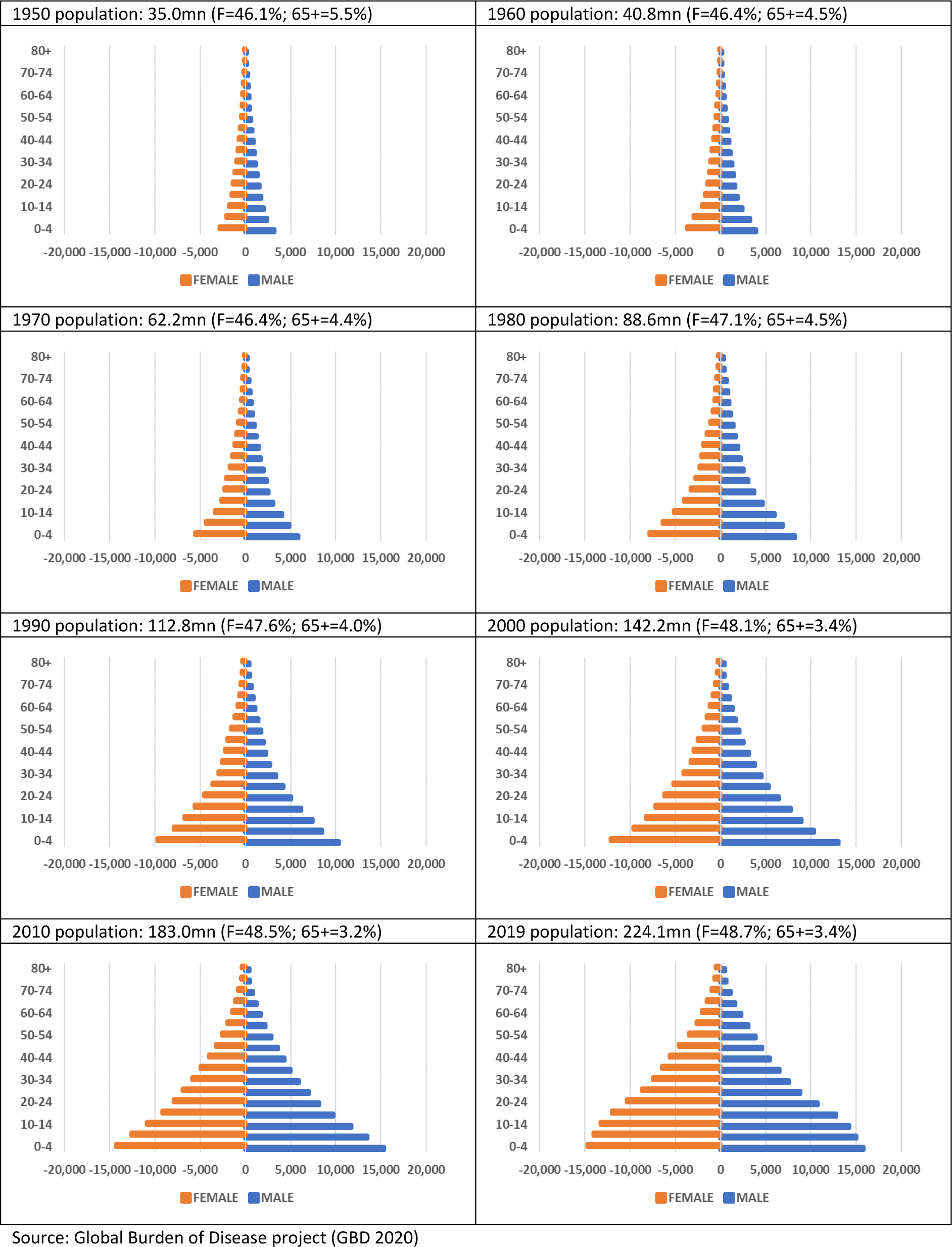
Pakistan population structure & population growth (1950-2019)

**Figure B.2.**
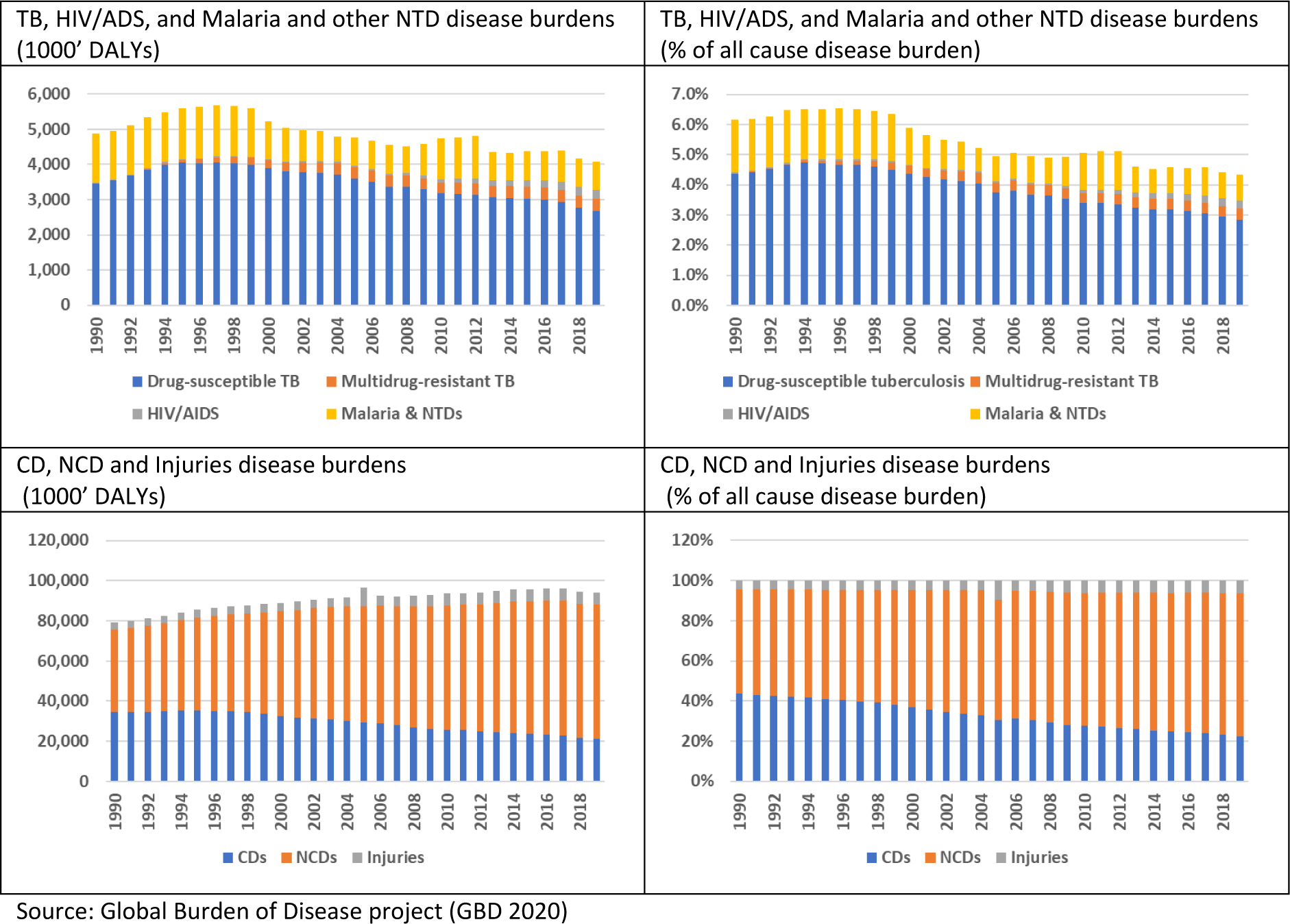
Structure and evolution of Pakistan disease burdens 1990-2019.

**Figure B.3.**
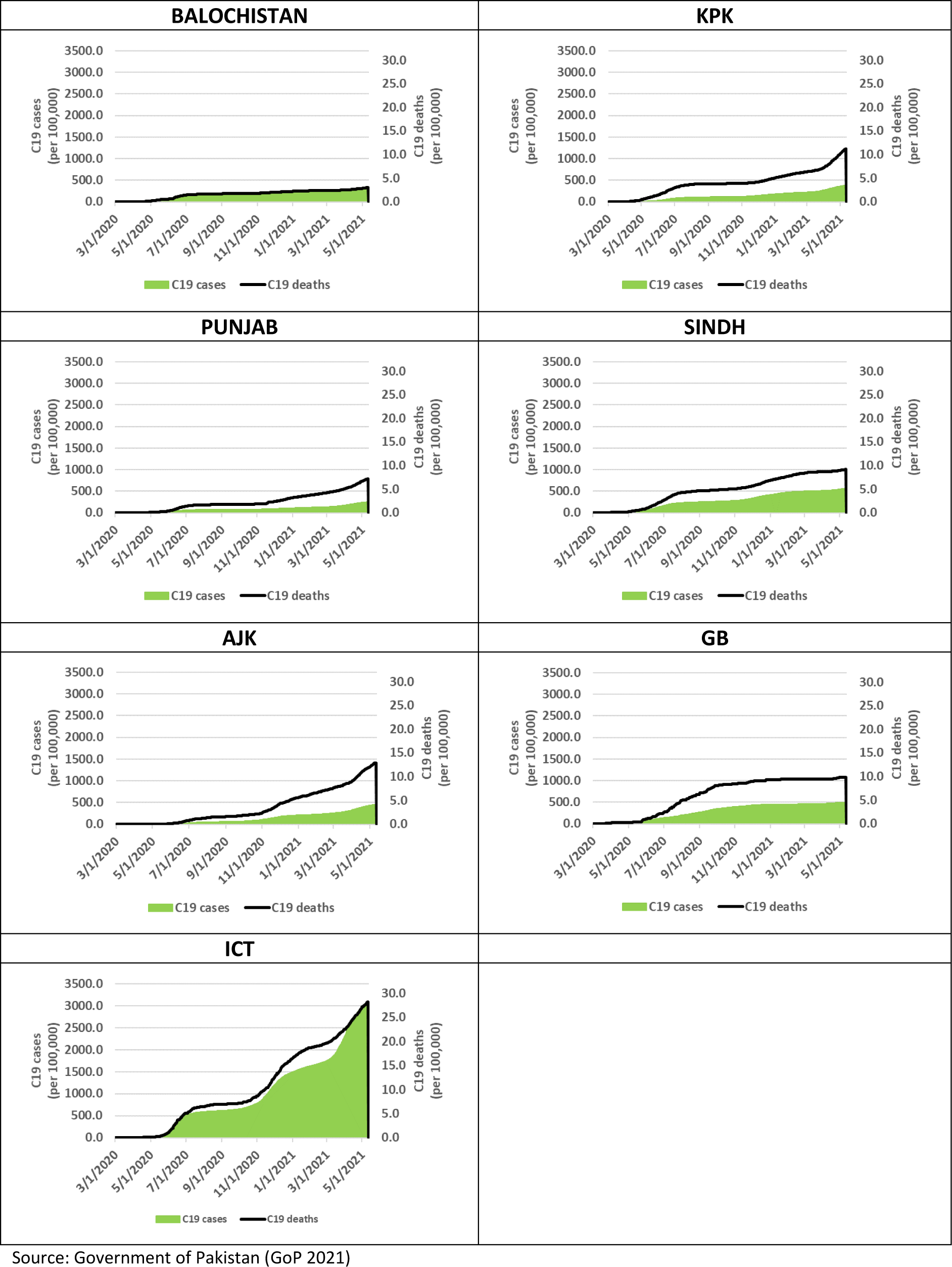
Pakistan C19 clinical outcomes by Province & Territory 1. March 2020 – 11. May 2021 (per 100,000 population)

**Figure B.4.**
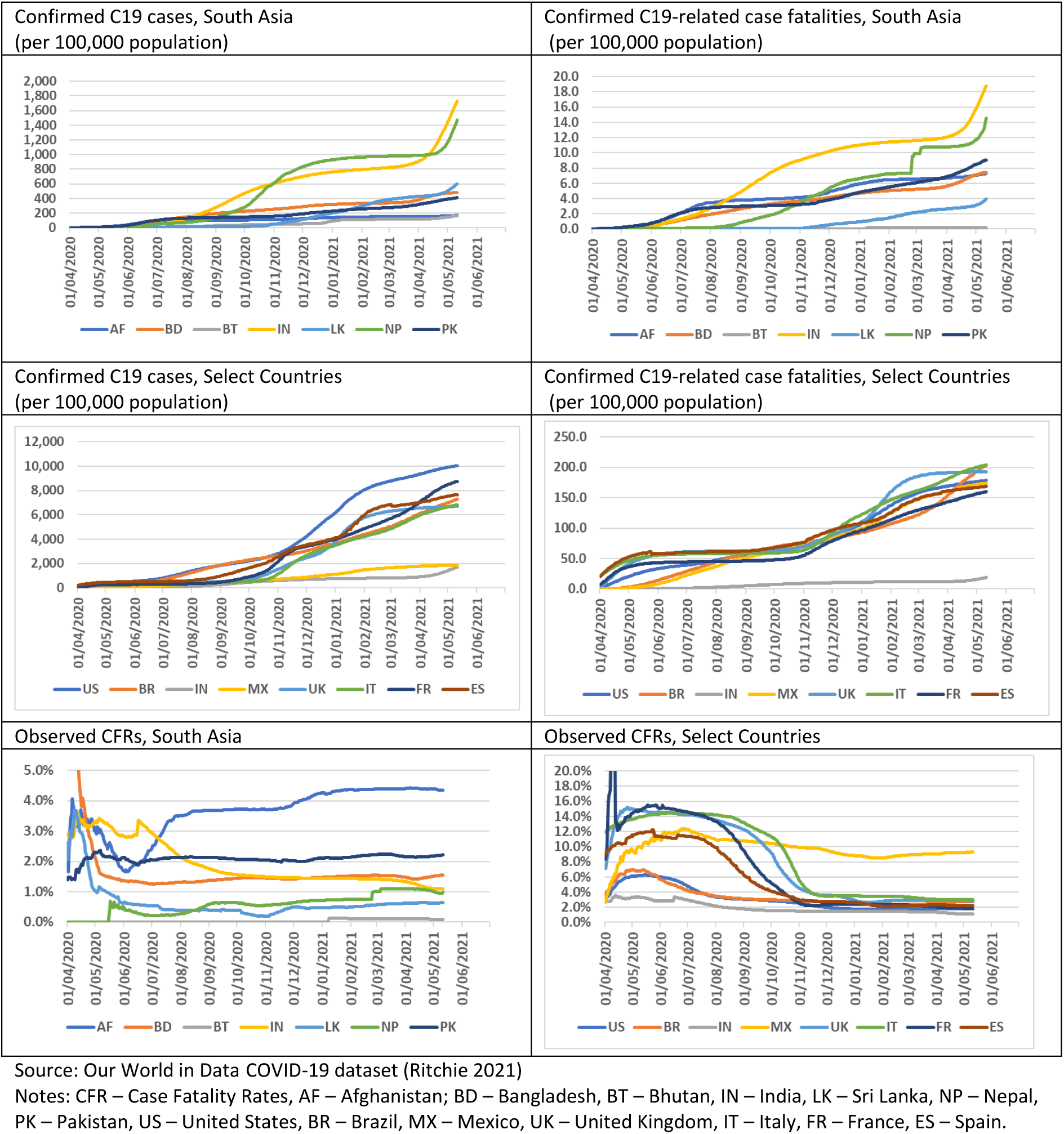
South Asia & select country C19 clinical outcomes 1. March 2020 – 11. May 2021.

**Table B.1.**
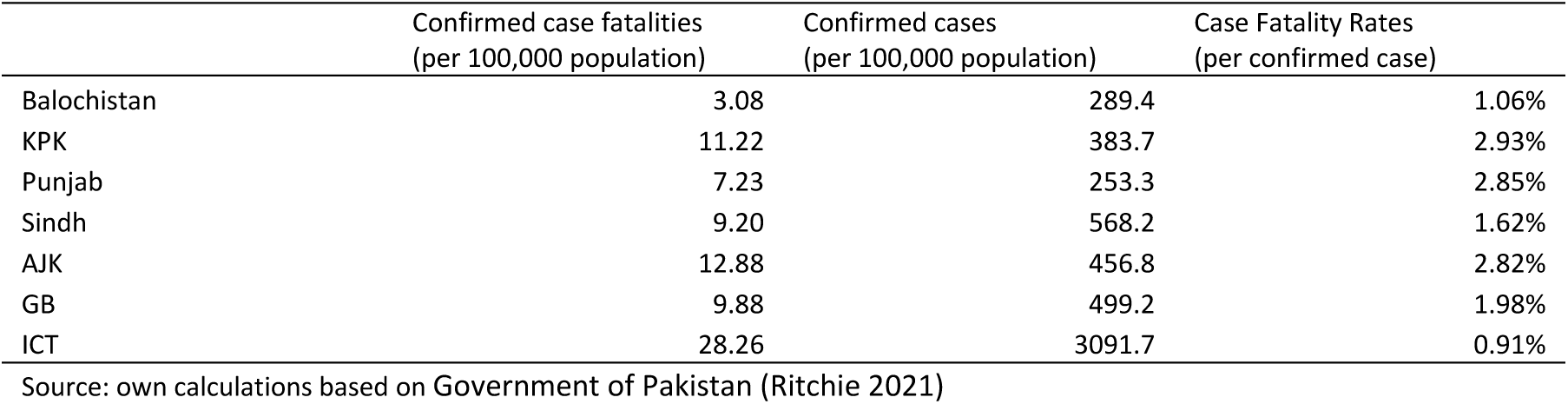
Confirmed C19 cases, case fatalities and CFRs (by 11 May 2021)

## Appendix C. Macroeconomic methods background

### C.1. CGE model parameters

in terms of CGE model parameterization and macro-closure, household demand is governed by a set of Linear Expenditure Systems (LES); Production is specified as Constant Elasticity of Substitution (CES) functions of aggregate intermediate input demands (disaggregate commodity input demands are determined by Leontief specifications) and aggregate factor input demands (disaggregate factor input demands are also determined by CES specifications) with standard elasticity values for the top-level production specifications (0.8) and the bottom-level factor input demand specifications (0.6); Trade between domestic and foreign agents is specified as a function of relative prices (determined by the real exchange rate), based on Armington CES specifications on the import side and Constant Elasticity of Transformation (CET) specifications on the export side. Standard trade elasticity values were applied on the import side (1.2) and on the export side (1.5).

### C.2. CGE model macro closure

Our macroeconomic model closure specifies the GDP deflator as price numeraire, i.e. it is kept fixed at the counterfactual growth path. Furthermore, our counterfactual growth path is simulated with a standard neoclassical model closure involving (1) price clearing of all goods and factor markets, (2) real exchange rate clearing the Balance of payments, and (3) investment is savings-driven. In addition, the counterfactual growth path are simulated with balanced macro-closure which ensures that the government consumption share of domestic absorption is fixed along the counterfactual growth path. The unmitigated C19 baseline scenario is, similarly, simulated with a standard neoclassical macro closure, but government consumption is ‘fixed’ at the counterfactual growth path (in terms of its own price deflator) and the simulations also involve a standard revenue-neutral government budget closure (household direct taxes specified as endogenous, in order to compensate for shocks to government expenditures).

The macroeconomic closures of the policy scenarios are slightly different involving separate closures for the individual BC-NPI, SC-NPI and HQ-NPI scenario components. The SC-NPI and HQ-NPI scenario components are simulated with a standard neoclassical model closure, whereas the BC-NPI component of the scenarios are simulated with a modified neoclassical model closure where factor demands are fixed (and reduced) in non-essential production sectors (with sector-specific factor prices clearing sector-specific factor demands), while uniformly varying factor prices continue to clear factor markets among essential production sectors. Otherwise, all policy scenario NPI components are simulated with (1) price clearing of all goods markets, (2) real exchange rate clearing the Balance of payments, and (3) investment is savings-driven; And they also keep government consumption ‘fixed’ at the counterfactual growth path and rely on a standard revenue-neutral government budget closure.

### C.3 Macroeconomic literature review

The macroeconomic C19 literature generally focuses on C19 as a health shock with economic repercussions (1) directly via health-related impacts and (2) indirectly via private and public mitigating actions. Broadly speaking, the literature distinguishes between direct and indirect economic impacts, supply and demand shocks, private behaviour and public policy responses, emergency and recovery policy measures, pharmaceutical interventions (PIs) and NPIs, mitigation and suppression interventions, and a host of other focus areas.

Research into the economics of C19 has, to a large degree, focused more on high-income countries (HICs) than on low- and middle-income countries (LMICs). However, some comprehensive monographs have emerged covering both global, HIC and LMIC perspectives (Baldwin & di Mauro 2020), and economists from international institutions have attempted to summarize the limitations which LMICs are facing in terms of structural limitations and potential policy options going forward (Loaysa & Pennings 2020).

A number of macroeconomic studies have emerged which focuses on combined epidemiological and macroeconomic modelling of the Covid-19 illness and aggregate Covid-19 NPI policy shocks. The group of aggregate Dynamic Stochastic General Equilibrium (DSGE) model studies, with endogenous epidemiological models, is focussed on aggregate social planning problems and optimal control strategies. In contrast, simple single-country fixed-price general equilibrium Multiplier Model (MM) studies, and more elaborate single-country and global Computable General Equilibrium (CGE) studies, are focussed on more detailed economic burden and NPI policy assessments; and, for one of the CGE studies (Keogh-Brown et al 2020) and one epidemiological-MM type model framework (Haw et al 2020a, 2020b), focus is also on detailed epidemiological modelling of clinical health outcomes. The two groups of macroeconomic modelling studies are described in the following.

#### Macroeconomic Social Planning DSGE studies

The group of aggregate DSGE intertemporal model studies employ simple but fully integrated SIR-type epidemiological models (Atkeson 2020a, 2020b). They also specify explicit social planning problems and use these to measure optimal lockdown strategies for minimizing macroeconomic and social welfare costs (Alvarez, Argente & Lippi 2020; Eichenbaum, Trebelo & Trabandt 2020; Jones, Philippon & Venkateswaran 2020a, 2020b).

The DSGE studies’ optimal strategies, typically, turn out to, essentially, analyse optimal strategies for keeping case fatalities low. Hence, their social planning problems, which have an infinite time horizon, are typically dominated by case fatalities’ loss of lifetime earnings, compared to which recovered individuals’ morbidity-related loss of earnings are small.

The highly aggregated nature of DSGE model applications has several drawbacks. Hence, most DSGE studies rely on simplified economic specifications, including limited sector-level detail and simplified economic production specifications with labour as only production factor (Alvarez, Argente & Lippi 2020; Eichenbaum, Trebelo & Trabandt 2020; Jones, Philippon & Venkateswaran 2020a, 2020b), and simplistic SIR epidemiological model specifications (Atkeson 2020a, 2020b). While the integrated epidemiological model, in one case, accounts for variation in ‘meeting’ rates between work, consumption (presumably a proxy for home contacts), and other meetings (Eichenbaum, Trebelo & Trabandt 2020), they generally lack nuanced socioeconomic policy scenario specifications of lockdown strategies (typically focusing on the BC-NPI aspect of lockdown strategies, but missing key distinctions between essential and non-essential sectors, and also missing nuances of school closures, home quarantines, etc.)

The DSGE studies also typically lack epidemiological modelling of age-specific heterogeneity both in terms of contact patterns and clinical health outcomes, and private agents are assumed homogenous thereby disregarding the potentially catastrophic (distributional) income losses likely to be experienced by part of the population especially in LMICs. The optimal social planning problems also don’t account for other non-monetary (health) outcomes, e.g. mental health impacts of lockdown measures and timely hospital treatment of non-C19 patients, and their application of widely varying Value of Statistical Life (VSL) measures of mortality-related welfare losses presents an additional weakness, with magnitudes ranging from 20xGDPcap (Alvarez, Argente & Lippi 2020) to estimates in the order of 140xGDPcap (Jones, Philippon & Venkateswaran 2020a, 2020b).

A key strength of the social planning DSGE approach is that it has managed to explicitly model private prevention behaviour, as part of the social planner’s problem, and to model two key externalities related to private behaviour: (1) the infection externality (each individual only focussed on own protection without considering impact on society protection leading to undersupply of transmission-reducing behavioural change), and (2) the health system congestion externality (each individual not taking risk of HS congestion, and associated excess mortality, into account, again leading to undersupply of transmission-reducing behavioural change). The DSGE specifications of these externalities exclude altruistic or ethical private behaviour, but these studies do, in an important way, contribute to bringing attention to the potential undersupply of risk-reducing private behavioural change.

Another strength of the social planning DSGE studies is that they have managed to analyse important issues such as tele-working. Some of the social planning analyses suggest that optimal tele-working (if possible) could approach 40% (Jones, Philippon & Venkateswaran 2020), and, since the potential for tele-working has been measured to be approx. 37% of all (US) jobs (Dingel & Neyman 2020), this provides an indication of the significant scope for mitigating tele-working interventions to contribute to epidemic control in countries where this is an option. While opportunities for tele-working may be more limited for Pakistan and other LMICs, the above results are suggestive, and e.g. large-scall tele-schooling initiatives have also been rolled-out in Pakistan and elsewhere (see discussion below).

#### Macroeconomic Burden Assessment CGE studies

The CGE model studies of aggregate C19 shocks, currently in the public domain, includes application of a dynamically-recursive version of the “Standard Model” single-country CGE model framework (Keogh-Brown et al 2020), and applications of various global model frameworks, including the G-Cubed model developed by Warwick McKibbin and colleagues (McKibbin & Fernando 2020) and the ImpactECON Supply-Chain Model which is a modified version of the Global Trade Analysis Project (GTAP) model (Walmsley, Rose & Wei 2020).

While the GTAP-inspired ImpactECON Supply-Chain Model is a standard CGE model, the G-Cubed model is a hybrid DSGE/CGE model with aggregate intertemporal DSGE features but with an underlying multi-sector CGE model which solves for annual equilibria over a very long time horizon (currently 2015-2100). The intertemporal features of the G-Cubed model, including long-term budget constraints, makes it suitable for long-term policy analyses, while the CGE features allows for appropriate short-to medium-term analyses. The G-Cubed model, is, however, a bespoke and very data-intensive model, and therefore not useful as a basis for country-specific applications.

Among the global studies, Walmsley et al employs a fairly simplistic specification of shocks, including only mandatory BC-NPIs of one and four month durations, while McKibbin and Fernando, in spite of being published very early (29 February 2020) and using scant data including 2003 SARS data from China and rough indicators for inferring country-specific shocks, goes much further and designs global sets of sophisticated country-specific scenarios involving direct and indirect health-related shocks (morbidity and mortality-related labour force impacts), equity risk premia, sector-specific (transport) costs of production, and consumer preference impacts (public health costs of clinical health outcomes are, however, not inferred, and therefore not analysed, and lockdown measures are not analysed either presumably due to the early publication date).

While neither the health-related impact focussed McKibbin-study or the NPI-focussed Walmsley-study have managed to successfully predict (or properly model the full range of mechanisms underlying) economic C19 impacts over 2020, it is interesting to note (1) the McKibbin-study findings that “Hong Kong Flu”- and “Spanish Flu”-type pandemics would lead to health-related costs equivalent to respectively 2% and 8% declines in global GDP ($2.3 & $9.2 trillion), and (2) the Walmsley-study predictions of four months of business closures leading to a 21.6% decline of U.S. GDP ($4.6 trillion) and a 23.0% drop in employment (36.1 million unemployed workers). While neither of these studies’ health-related or NPI intervention scenarios matched what actually happened during 2020, they do suggest possible orders of magnitudes of (1) health-related costs if no NPI interventions had been undertaken, and (2) NPI costs if comprehensive BC-NPIs had been implemented without attempts to introduce either substitute NPIs (e.g. SC-NPIs, HQ-NPIs, and GG-NPIs) or complementary NPI interventions to lower BC-NPI impacts (e.g. tele-working). In this sense, these early global studies underline the critically important point, that, while business closures can affect contact numbers at work, BC-NPIs are extremely costly to society, and, if alternative NPI measures aren’t effectively implemented and managed (e.g. through communication campaigns from trusted authorities), epidemic control may, in the absence of effective PIs, be very costly to society.

Using the same “Standard Model” single-country methodology as the current study (see Methods section below), a UK single-country study analysed C19 lockdown scenarios with a broader scope of NPI interventions involving both BC-NPIs, SC-NPIs, and HQ-NPI components (Keogh-Brown et al 2020). The UK CGE model was specified to mimic the detailed epidemiological predictions of the widely recognized Imperial College model for the UK, including modelling of their preferred intermittent 2-month lockdown/1-month opening-up scenario (Ferguson, Laydon et al. 2020). The business closure component was, however, not analysed with complementary interventions, e.g. tele-working, and simulated magnitudes of economic costs of NPIs, necessary to control the UK epidemic, were therefore on a par with the global Walmsley-study (essentially, predicted economic impacts could be viewed as a worst-case scenario).

A key strength of the single country CGE framework, in a health context, is its ability to accommodate exogenous epidemiological model predictions, and thereby allow for model-consistent public health assessments of actual monetary policy impacts and economic burdens (including health system costs and labour market impacts), alongside clinical health burdens, for individual illnesses (Jensen et al 2019a) and national epidemics (Keogh-Brown et al 2020), but without making assumptions about government preferences. Hence, the focus on analysing policy scenarios, rather than specifying social planning problems, allow decisions on trade-offs to be left up to policy makers – and, by the same token, policymakers are free to apply their own preferences when they weigh relative economic and clinical health impacts from various NPI policy scenarios.

Turning to the C19 applications of MM models, also known as fixed-price general equilibrium models, they have been applied, mostly by IFPRI-researchers and their peers, to assess potential economic impacts of C19 epidemics across a range of developing countries including Ghana (Amewu et al 2020), Nigeria (Andam et al 2020), Ethiopia (Aragie, Taffesse & Thurlow 2020), South Africa (Arndt et al 2020), Myanmar (Diao et al 2020), and Indonesia (Pradesha et al 2020). Within the MM model literature, a distinction is typically drawn between (1) Leontieff MM models (endogenous accounts only including production activity and retail commodity accounts), and (2) SAM MM models (endogenous accounts typically also including factor, enterprise and private household accounts, in order to allow for endogenous income-feedback loops via private consumption). The key motivation for applying the MM methodology, in the context of the C19 epidemic, seems to have been to derive very short-term impacts given that the shocks analysed are “…working through the economy in weeks or months…” (Diao et al 2020), and the authors further underline their point by saying that “(T)he SAM multiplier model is a simulation tool suited to measuring short-term direct and indirect economy-wide impacts of unanticipated, rapid-onset economic shocks, such as COVID-19” (ibid.) Another study adds that it is the unconstrained MM methodology which is applied (Andam et al 2020), something which seems to be implicitly assumed in the other IFPRI-studies.

In terms of MM model closure specifications, one IFPRI-study simply states that “(T)he SAM-multiplier model deployed treats the elements of final demand (household consumption, investment, government expenditure, and exports) as exogenous variables that can be ‘shocked’” without further motivation (Arndt et al 2020), while another explains that, beyond other final demand components, the model “…also treats household consumption demand as exogenous, meaning that an income shock does not result in subsequent rounds of consumption demand and income feedbacks. This avoids over-estimating multiplier effects…” (Andam et al 2020). As noted above, this is a non-standard SAM MM model closure. It can also be noted that economic multipliers, derived from MM models, have no time dimension. The multiplier formula *M* = (*I* − *A*)^−1^ = *A* + *A*^2^ + ⋯ + *A*^*n*^ + ⋯ indicates that (Leotieff and SAM) multipliers, in fact, are the result of an infinite number of economic feedback-loops, but since these loops do not have a time dimension, it is not possible to attach a specific time dimension to the multipliers. While some studies explicitly states that exclusion of the income-feedback loop is made to avoid overestimating multipliers, including (Amewu et al 2020) referencing (Haggblade and Hazell 1989), there also seems to be an implicit assumption that exclusion of the income-feedback loop can motivate interpreting multipliers as very-short term. This is however not the case. The IFPRI-studies’ model closure implies that Leontieff multipliers, with their narrow focus on intermediate demand-feedback loops, are driving their multiplier results, and since these multipliers are also the results of infinite numbers of iterations, it is still not clear that the IFPRI-studies’ multipliers can appropriately be given a very short-term interpretation.

Another weakness of the IFPRI MM model approach is that MM models assume linear production technologies and fixed prices (Andam et al 2020), and, while it is argued that CGE models are inappropriate for analyzing C19 lockdowns “…where trade and production have ceased due to government directives, rather than by market forces…” and that MM models, by default, are preferred (ibid.), the implicit factor market closure for the unconstrained MM model, i.e. excess supplies of factor resources (including capital), is unrealistic for most LMICs. In fact, while many of the IFPRI-studies was admittedly written during the early stages of the pandemic and therefore hasn’t been able to benefit from knowing how LMIC policy responses have unfolded during 2020, the reality of C19 policy responses in most LMICs, perhaps with the exception of South Africa, has been, that hard business lockdowns have been avoided, or, if attempted, they have quickly been substituted for softer ‘smart’ lockdown strategies. This switch in strategy towards smart lockdowns was made, exactly to avoid the catastrophic poor informal sector household income loss scenarios which several IFPRI-studies points out would have been the result of such lockdowns. As such, the IFPRI MM model impacts, same as the excessive CGE model impacts referred to above, can be considered as worst-case NPI costs.

Also, the fact that hard lockdowns has been the exception in LMICs, invalidates the key argument that “…trade and production have ceased…” Hence, as argued above, most country responses to C19 have gone to great pain to focus on reducing other contact numbers before resorting to BC-NPIs, and, when resorting to “last resort” BC-NPIs, they have typically been partial and targeted at hospitality sectors with particularly high contact numbers. With this in mind, and also considering that LMICs, including Pakistan, have focused on smart lockdowns in order to allow their (informal) service sector to continue to operate, it is reasonable to assume that price adjustment has continued to play a key role in resource allocation in most LMICs, and that CGE models therefore remains an appropriate tool even in the face of multiple simultaneous demand-side shocks and supply-side NPI policy restrictions.

Perhaps the greatest weakness of IFPRIs unconstrained MM model applications is, that MM models are inherently demand-driven. The implicit MM model closure implicitly assumes that final demand is the restricting factor, while, as mentioned above, supply-side resources, including labour and capital factor supplies, are freely available to adjust. While uncertainty, created by national C19 epidemics, are bound to have impacted domestic final demand components, this model closure is not consistent with modelling of, essentially, supply-side lockdown interventions which closes factor capital equipment down and reduces effective labour force participation rates.

In spite of the above-mentioned drawbacks, it should be noted that a key strength of the IFPRI MM Model applications is that they build on economywide SAM data sets, and, while economic data from LMICs has become much more accessible with the proliferation of economic survey data since the early 1970s (Thorbecke 2000), SAM data sets continue to provide almost unrivalled access to economy-wide policy analysis in many developing countries. And, with the simplicity of the SAM MM methodology, this has allowed for quick access to (simple, but timely and policy-relevant) policy analyses to inform policy makers in LMICs.

Finally, it should be mentioned that Imperial College has established an epidemiological-MM type model framework – a framework which is related to the macroeconomic social planning DSGE studies, discussed above, since this framework also consists of an epidemiological model coupled with a macroeconomic module. These frameworks are linked together by (1) spillovers from the epidemiological model to the macroeconomic model, and (2) a social planner objective function which, in this case, is real GDP (Haw et al 2020a, 2020b). The Imperial framework is, however, distinguished, from the DSGE model frameworks, by a much more sophisticated approach to epidemiological modelling, but also by a rudimentary macroeconomic module, consisting, purely, of fixed sector-specific production vectors, which are scaled by sector-specific BC-NPI lockdown shares in order to compute linear variations in sector-level GDP, but which does not involve economic behaviour.

## Footnotes

1 Throughout January-February 2021, WHO reported no community transmission of B117 (WHO 2021d), which is surprising given that it emerged as the dominant strain in Punjabi urban centres including the metropolis of Lahore in early-March 2021 (SAMAA 2021).

2 Based on the functional relationship between R0, vaccine efficacy (ε) and required coverage rates for herd immunity (COVER), COVER = [1-(1/R0)]/ε (Anderson et al 2020).

3 The now dominant B117 strain reportedly has 50-74% higher transmissibility (Davies, Barnard et al 2020), and other variants of concern, including B1351 and P1, which had been verified in Pakistan in early-May 2021, (WHO 2021e), reportedly has similarly elevated transmissibility (Faria et al 2021).

4 See appendix B for details

5 Mexico had only recorded 2,958 confirmed cases per 100,000 on 14. October 2021. However, the low confirmed case count is likely to be significantly underestimated, as confirmed by the high case fatality numbers (224 per 100,000) which themselves are reported to be underestimated by 60% (Reuters 2021h).

6 No peer reviewed studies have so far confirmed the 79% efficacy level for BBIBP-CorV.

7 The slow roll-out prompted health authorities to post warnings to HCWs, including the Sindh Directorate of Health Services who, in a letter dated 24 February 2021, warned HCWs that they could face “strict disciplinary action” if they didn’t get vaccinated, but HCWs seem to have remained reluctant (HPW 2021).

8 Further delays of COVAX deliveries were reported end-of-March 2021, due to redirection of vaccine deliveries, by Serum Institute of India, in the context of rising Indian transmission (Reuters 2021j, Reuters 2021k). The coming COVAX-related vaccine deliveries, including shipment of 2.4mn ChAdOx1 doses in June 2021, are instead expected to arrive from South Korea (Reuters 2021n).

9 We had to exclude the two autonomous territories of Gilgit-Baltistan (GB) and Azad Jammu & Kashmir (AJK), and the federal Islamabad Capital Territory (ICT) from our household disaggregation, since the 2015-16 HIICS survey only included survey data for the four main provinces of Pakistan (PBS 2017). However, since the four provinces is estimated to account for 94.7% of the total Pakistan population in 2020 (WorldPop 2020), our analyses are likely to provide a good approximation to Pakistan-wide impacts.

10 The derivation of Pakistan-specific R0 estimates were, in some cases, based on alternative approaches to epidemiological modelling including standard SEIR-model frameworks (Ullah & Khan 2020), and other multi-compartmental transmission model frameworks (Ali, Imran & Khan 2020).

